# Comparative sensitivity evaluation for 122 CE-marked SARS-CoV-2 antigen rapid tests

**DOI:** 10.1101/2021.05.11.21257016

**Authors:** Heinrich Scheiblauer, Angela Filomena, Andreas Nitsche, Andreas Puyskens, Victor M Corman, Christian Drosten, Katrin Zwirglmaier, Constanze Lange, Petra Emmerich, Michael Müller, Olivia Knauer, C Micha Nübling

**Affiliations:** Paul-Ehrlich-Institute, Paul-Ehrlich-Str. 51-59, D-63225 Langen; Robert Koch-Institute, Seestrasse 10, D-13353 Berlin; Institute of Virology, Charite, Chariteplatz 1, D-10117 Berlin; Bundeswehr Institute of Microbiology, Neuherbergstr 11, D-80937 Munich; LADR GmbH, Lauenburger Str. 67, D-21502 Geesthacht; Bernhard-Nocht Institute, Dep.Virology, Bernhard-Nocht Str. 74, D-20359 Hamburg; MVZ Labor 28 GmbH, Mecklenburgische Str. 2, D-14197 Berlin

## Abstract

**Objective:** Independent evaluation of the sensitivity of CE-marked SARS-CoV-2 antigen rapid diagnostic tests (Ag RDT) offered in Germany.

**Method:** The sensitivity of 122 Ag RDT was adressed using a common evaluation panel. Minimum sensitivity of 75% for panel members with CT<25 was used for differentiation of devices eligible for reimbursement in in the German healthcare system.

**Results:** The sensitivity of different SARS-CoV-2 Ag RDT varied over a wide range. The sensitivity limit of 75% for panel members with CT <25 was met by 96 of the 122 tests evaluated; 26 tests exhibited lower sensitivity, few of which were completely failing. Some devices exhibited high sensitivity, e.g. 100% for CT<30.

**Conclusion:** This comparative evaluation succeeded to distinguish less sensitive from better performing Ag RDT. Most of the Ag RDT evaluated appear to be suitable for fast identification of acute infections associated with high viral loads. Market access of SARS-CoV-2 Ag RDT should be based on minimal requirements for sensitivity and specificity.

## Introduction

A large number of antigen-detecting rapid diagnostic tests (Ag RDTs) for SARS-CoV-2 are available on the European market, both for professional use and as self-tests. Rapid tests are based on lateral flow immunochromatography using antibodies against SARS-CoV-2 proteins (antigens), present in respiratory tract specimens. By far most Ag RDTs target the viral nucleoprotein (N), only very few assays work with spike protein (S) detection. Viral variants of concern (VOC) have been described mainly for the S gene, leaving the vast majority of SARS-CoV-2 Ag RDT unaffected; however, the few SARS-CoV-2 Ag RDT based on spike protein detection should be checked at regular intervals for potential deficiencies. While PCR is still the gold standard for virus detection, there is an increasing evidence that infectivity by respiratory secretions correlates with high viral loads present in the early phase of infection, e.g. before and after (0-10 days) onset of symptoms. Thus Ag RDTs allow rapid identification of acutely infected and potentially infectious individuals facilitating fast decisions on containment of virus spread, patient care, isolation and contact tracing. Furthermore, Ag RDTs may save limited reagents of more sensitive molecular diagnostics to serve other diagnostic needs, e.g. disease management or confirmation of Ag RDT reactive results.

In the European Union (EU), regulatory requirements for SARS-CoV-2 *in vitro* diagnostic medical devices (IVD) are defined by the IVD Directive 98/79/EC (IVDD) and have to be addressed by the manufacturer prior to access to the EU Common Market. However, certification (CE-marking) of SARS-CoV-2 diagnostics is currently done solely by the manufacturer (self-certification), without third party intervention. The exception are SARS-CoV-2 self-tests, where a notified body has to assess the lay person studies. However, due to the urgency in the Corona situation a national derogation can be agreed by the national Competent Authority, e. g. based on the performance of an identical test for professional use. From May 2022 the IVDD will be replaced by the IVD Regulation (EU) 2017/746 (IVDR) where a risk-based classification of IVDs is the basis for the scrutiny of their assessment (1). SARS-CoV-2 IVD will belong to the high-risk devices (class D) under the IVDR, requiring a Notified Body both for certification of the manufacturer`s quality management system and for assessment of the technical documentation of the device. Furthermore, EU reference laboratories (EURL) will be responsible for independent laboratory testing of class D devices to verify performance features and to assure batch to batch consistency.

However, at the time being independent evaluations of SARS-CoV-2 Ag RDT that allow conclusions on their performance are largely missing.

In the current situation with absence of strict regulatory requirements for most SARS-CoV-2 IVD, the German Ministry of Health decided to link the reimbursement of SARS-CoV-2 Ag RDT to provision of evidence of essential quality features of these assays. This evidence consisted of two parts: 1) provision by the manufacturer of evidence for compliance with minimum criteria, and 2) successful independent laboratory evaluation. Minimum criteria for sensitivity and specificity were jointly defined by Paul-Ehrlich-Institut (PEI) and Robert Koch Institut (RKI), two governmental authorities in Germany (2). Manufacturers or distributors of SARS-CoV-2 Ag RDT document for the respective SARS-CoV-2 Ag RDT compliance with these minimum criteria before the device can be listed as eligible for reimbursement on a dedicated webpage of Bundesinstitut für Arzneimittel und Medizinprodukte (BfArM), another governmental authority (3).

Devices were selected from the BfArM list for the comparative evaluation performed by PEI/RKI. The aim of this comparative evaluation was to both determine the “state of art” sensitivity of proficient devices and identify devices not reaching the minimum sensitivity level. Subsequently, devices with sensitivity below “state of the art” were removed from the BfArM list while all devices with successful evaluation outcome were published on PEI webpage (4). In the meantime more than 120 SARS-CoV-2 Ag-RDT have been evaluated in direct comparison using common SARS-CoV-2 specimens. The outcome and conclusions of this study are summarized in this manuscript.

## Materials and Methods

### Evaluation panel

Detailed characterization of the evaluation panel has been described by Puyskens et al. in the tandem publication to this study. In short, pools from nasopharyngeal and oropharyngeal swabs were prepared as random mixtures obtained from up to 10 swabs. While dry swabs were directly eluted in phosphate buffered saline (PBS), the residual amount virus transport media (VTM) contained in moist swabs was diluted in PBS. Care was taken not to use VTM containing the protein denaturing component guanidinium.

Individual pools are composed of samples with similar SARS-CoV-2 concentrations, expressed as cycle threshold (CT) values of semiquantitative PCR. In total 50 different pools were defined as members of the evaluation panel and stored as 500 µl aliquots at -80°C. The CT of each panel member was determined by PCR, and the putative number of RNA copies calculated with the aid of the reference preparations distributed by the German external quality assessment (EQA) provider INSTAND e. V. (5). Furthermore, presence of infectious virus detectable by propagation in Vero cell culture was determined for the individual pools.

The whole evaluation panel may be subdivided into three subgroups: panel members, which are characterized by very high (CT 17-25; 18 pools), high (CT 25-30; 23 pools) or moderate (CT 30-36; 9 pools) viral load. During the comparative evaluation 4 panel members of the original panel had to be replaced, resulting in a slight shift of subgroups composition in the resulting panel version 2: 17 pools covering the CT-range 17-25, 23 pools the CT range 25-30 and 10 pools the CT range 30-36.

### Antigen stability

Real-time antigen stability in panel members was investigated using quantitative SARS-CoV-2 ELISA Lumipulse G SARS-CoV-2 Ag (Fujirebio Inc., Shinjuku-ku, Tokyo, Japan). Panel members were tested after initial thawing and throughout one week incubation at 4°C. Furthermore, potential impact of additional freeze/thaw cycle was addressed.

### Comparative evaluation

In the beginning of the comparative evaluation, laboratories participating in the comparative evaluation included the Robert Koch-Institute, the Paul-Ehrlich-Institute, the Nationales Konsiliarlaboratorium für Coronaviren (Charité), the Bundeswehr Institute of Microbiology, the Bernhard-Nocht-Institut für Tropenmedizin, and laboratories of the association ALM (Akkreditierte Labors in der Medizin). At a later stage, because of the massively increasing work load, the evaluation was continued mainly by PEI and RKI. Panels were shipped on dry ice and, once thawed, 50 µl aliquots were prepared, kept at 4°C and used within 5 days, without further freeze / thaw step. For each Ag RDT and panel member, the 50 µl aliquot was completely absorbed using the specimen collection device, e.g. swab, provided with the respective test. The swabs were then eluted in the test-specific buffer, strictly following the respective instructions for use (IFU). After applying the sample/buffer solution onto the test cassette and incubation, visual read out of control and target lines was done independently by two lab technicians, with potential discrepant results preliminarily interpreted as “equivocal”. In favor of the tests evaluated, both reactive and equivocal results for the target line were eventually scored as positive. At PEI the test cassettes were immediately scanned using BLOTrix Reader R2L (BioSciTec GmbH) and analysed with BLOTrix 4 Cubos (B4C) software (BioSciTec GmbH), at other evaluation sites the test results were documented by photographs. Some tests were provided with reading instruments and read as per instruction manual provided.

Tests were selected from original manufacturers, as far as this information was available. Often duplicate version of the very same tests are marketed under a new test name, new manufacturer or different distributor. Repeat testing of duplicates was avoided as far as possible in order to cope with the already huge variety of different tests placed on the EU Common Market.

## Results

### Characterization of the evaluation panel

Panel members spanned the CT range between 17 and 36. A specimen with an assigned SARS-CoV-2 RNA concentration of 10^6^ RNA copies/ml provided by INSTAND corresponded to the CT value of 25. Assuming that a CT difference of 1 corresponds to a concentration factor of 2 and taking into account that the individual panel members cover a CT range from 17 to 36, the SARS-CoV-2 RNA amounts in the panel cover a concentration range from >10^8^ to <10^3^ copies / ml, respectively. The CT values of 20 or 30 would than correspond to approximate SARS-CoV-2 RNA concentrations of 3 × 10^7^ or 3 × 10^4^ copies / ml, respectively.

SARS-CoV-2 propagation in cell culture resulted in positive results for several of the low CT / high titre specimens, indicating presence of infectious virus despite the various preparation steps (more details in the tandem publication of Puyskens et al).

Stability of the analyte SARS-CoV-2 antigen in all panel members was studied under different conditions using a quantitative ELISA. While additional freeze/thaw steps had negative effect on analyte stability, there was no significant impact on the antigen content after up to 7 days storage at 4°C of the liquid 50 µl aliquots (data not shown). From one 500 µl thawed aliquot, nine to ten 50 µl aliquots were immediately filled and used within 5 days for the corresponding number of 9-10 test kits, ensuring no stability issues.

### Comparative evaluation

122 SARS-CoV-2 rapid tests were evaluated in direct comparison using the evaluation panel, with only few differences between the closely related panel versions 1 and 2. For acceptable Ag RDT performance a minimum sensitivity of 75% for the panel member subgroup with very high SARS-CoV-2 concentration (CT<25, viral load around 10^6^ SARS-CoV-2 RNA/ml and higher) was defined. This criterion corresponds to the detection of at least 14 of 18 subgroup positives in panel version 1 (18 members with CT<25), or 13 of 17 (17 members with CT<25 in panel version 2), respectively.

Of the 122 SARS-CoV-2 Ag RDT evaluated, 96 tests (78.7%) had a sensitivity of >75% for panel members with high viral loads (CT<25; Table 1), and 26 tests (21.3%) were of lower sensitivity not meeting the sensitivity criterion (Table 2). Of the 96 tests meeting the sensitivity limit, 58 (60.4%) detected all panel members of the subgroup with CT< 25 (100% subgroup sensitivity), and another 17 tests (17.7%) exhibited a respective subgroup sensitivity of >90%. In addition, 19 tests (19.8%) showed a sensitivity of >75% even in the CT range 25-30.

**Table 1.** Comparative evaluation results of SARS-CoV-2 antigen RDT passing the sensitivity criteria. (in alphabetical order of manufacturers)

| No. | Manufacturer | Test name | Sensitivity |  |  |  |
| --- | --- | --- | --- | --- | --- | --- |
|  |  |  | CT <25 | CT 25-30 | CT >30 | CT 17-36 |
| 1 | Abbott Rapid Diagnostics Jena GmbH | Panbio™COVID-19 Ag Rapid Test Device (NASOPHARYNGEAL) | 100,0% | 60,9% | 0,0% | 64,0% |
| 2 | ACON Biotech (Hangzhou) Co., Ltd | Flowflex SARS-CoV-2-Antigenschnelltest (Nasopharynxstupfer) | 94,1% | 4,3% | 0,0% | 34,0% |
| 3 | Aesku Diagnostics GmbH | Aesku Rapid SARS-CoV-2 Rapid Test | 82,4% | 17,4% | 0,0% | 36,0% |
| 4 | Affimedix | TestNOW® - COVID-19 Antigen | 100,0% | 47,8% | 0,0% | 58,0% |
| 5 | Amazing Biotech (Shanghai) Co., Ltd | CoroVisio Covid-19 Ag Versieglungsröhrchen Teststreifen (Kolloidales Gold) | 76,5% | 8,7% | 0,0% | 30,0% |
| 6 | Ameda Labordiagnostik GmbH | AMP Rapid Test SARS-CoV-2 Ag | 100,0% | 78,3% | 0,0% | 70,0% |
| 7 | AmonMed (Xiamen) Biotechnology Co., Ltd. | COVID-19 Antigen Rapid Test Kit (Colloidal Gold) | 100,0% | 87,0% | 30,0% | 80,0% |
| 8 | Anbio (Xiamen) Biotechnology Co., Ltd | Rapid Covid-19 Antigen Test (Colloidal Gold ) | 100,0% | 52,2% | 0,0% | 58,0% |
| 9 | Anhui Deepblue Medical Technology Co. , Ltd. | COVID-19 (SARS CoV-2) Antigen Test Kit (Colloidal Gold) | 100,0% | 39,1% | 0,0% | 52,0% |
| 10 | ASAN PHARM.CO.,LTD. | Asan Easy Test COVID-19 Ag | 100,0% | 69,6% | 0,0% | 66,0% |
| 11 | Atlas Link Technology Co.,Ltd. | Nova Test SARS-CoV-2 Antigen Rapid Test Kit | 100,0% | 60,9% | 0,0% | 62,0% |
| 12 | Avalun | Ksmart® SARS-COV2 Antigen Rapid Test | 94,1% | 13,0% | 0,0% | 38,0% |
| 13 | AXIOM Gesellschaft für Diagnostica und Biochemica mbH | Axiom Diagnostics COVID-19 Ag Schnelltest | 100,0% | 52,2% | 0,0% | 58,0% |
| 14 | Azure Biotech Inc. | Dia Sure Covid-19 Antigen Rapid Test Device (Nasopharyngeal/Oropharyngeal Swab) | 76,5% | 13,0% | 20,0% | 36,0% |
| 15 | Becton Dickinson | BD Veritor™ System for Rapid Detection of SARS-CoV-2 | 83,3% | 8,7% | 0,0% | 34,0% |
| 16 | Beijing Beier Bioengineering Co., Ltd. | Covid-19 Antigen Schnelltest | 77,8% | 0,0% | 0,0% | 28,0% |
| 17 | Beijing Hotgen Biotech Co., Ltd. | Novel Coronavirus 2019-nCoV Antigen Test (Colloidal gold) | 100,0% | 47,8% | 0,0% | 56,0% |
| 18 | Beijing Lepu Medical Technology Co., Ltd | SARS-CoV-2 Antigen Rapid Test Kit | 100,0% | 26,1% | 0,0% | 46,0% |
| 19 | Beijing Tigsun Diagnostics Co.,Ltd. | Tigsun COVID-19 Saliva Antigen Rapid Test | 100,0% | 87,0% | 30,0% | 80,0% |
| 20 | BIOMERICA Inc. | COVID-19-Antigen-Schnelltest (Nasopharyngeal-Abstrich) | 100,0% | 30,4% | 0,0% | 48,0% |
| 21 | BIONOTE | NowCheck® COVID-19 Ag Test | 100,0% | 65,2% | 0,0% | 66,0% |
| 22 | BioRepair GmbH | Covid 19 Antigen Schnelltest | 100,0% | 78,3% | 0,0% | 70,0% |
| 23 | BIOSYNEX SWISS SA | BIOSYNEX COVID-19 Ag BSS | 100,0% | 78,3% | 11,1% | 74,0% |
| 24 | BTNX, Inc. (Biotrend Chemikalien GmbH) | Rapid Response COVID-19 Rapid Test Device | 94,1% | 13,0% | 10,0% | 40,0% |
| 25 | Chil Tibbi Mal. San. Tic. Ltd. Şti | COVID-19 Antigen Schnell Test (Nasopharyngeal / Oropharyngeal Tupfer Kassette) | 100,0% | 60,9% | 0,0% | 62,0% |
| 26 | Core Technology Co., Ltd. | Canea COVID-19 Antigen Schnelltest | 88,2% | 26,1% | 0,0% | 42,0% |
| 27 | DNA Diagnostic A/S. | Covid-19 Antigen Detection Kit | 100,0% | 39,1% | 10,0% | 54,0% |
| 28 | Edinburgh Genetics Limited | Edinburgh Genetics ActivXpress+ COVID-19 Antigen Complete Testing Kit | 100,0% | 34,8% | 0,0% | 50,0% |
| 29 | Eurobio Scientific | EBS SARS-CoV-2 Ag Rapid Test | 94,1% | 34,8% | 0,0% | 48,0% |
| 30 | Fujirebio Inc. (Mast Diagnostica GmbH) | ESPLINE® SARS-CoV-2 | 100,0% | 21,7% | 0,0% | 46,0% |
| 31 | Genrui Biotech Inc. | Genrui SARS-CoV-2 Antigen Test Kit (Colloidal Gold) | 94,1% | 56,5% | 0,0% | 58,0% |
| 32 | GenSure Biotech Inc. | DIA-COVID® COVID-19 Ag Rapid Test Kit | 94,1% | 13,0% | 0,0% | 38,0% |
| 33 | Getein Biotech, Inc. | One Step Test for SARS-CoV-2 Antigen (Colloidal Gold) | 100,0% | 82,6% | 0,0% | 72,0% |
| 34 | Green Cross Medical Science Corp. (Weko Pharma GmbH) | Genedia W Covid-19 Ag | 83,3% | 8,7% | 0,0% | 34,0% |
| 35 | Guangdong Hecin Scientific,Inc. | 2019-nCoV Antigen Test Kit(colloidal gold method) | 82,4% | 13,0% | 0,0% | 34,0% |
| 36 | Guangdong Wesail Biotech Co., Ltd. | COVID-19 Ag Test Kit | 100,0% | 52,2% | 11,1% | 62,0% |
| 37 | Guangzhou Wondfo Biotech Co. Ltd | Wondfo SARS-CoV-2 Antigen Test (Lateral Flow Method) | 88,2% | 0,0% | 0,0% | 30,0% |
| 38 | Hangzhou Clongene Biotech Co., Ltd. | Clungene COVID-19 Antigen Rapid Test | 94,4% | 34,8% | 0,0% | 50,0% |
| 39 | Hangzhou Immuno Biotech Co.,Ltd. | IMMUNOBIO SARS-CoV-2 Antigen-Schnelltest (COVID-19 Ag) | 88,2% | 13,0% | 0,0% | 36,0% |
| 40 | Hangzhou Laihe Biotech Co., Ltd. (Lissner Qi GmbH) | Lyher Novel Coronavirus (COVID-19) Antigen Test Kit (Colloidal Gold) | 94,4% | 17,4% | 0,0% | 42,0% |
| 41 | Hangzhou Lysun Biotechnology Co., Ltd. | Lysun COVID-19 Antigen Rapid Test Device (Colloidal Gold) | 100,0% | 78,3% | 0,0% | 70,0% |
| 42 | Hangzhou Testsea Biotechnology Co., Ltd | Testsealabs® Rapid Test Kit COVID-19 Antigen Test Cassette | 100,0% | 47,8% | 0,0% | 56,0% |
|  |  |  | CT <25 | CT 25-30 | CT >30 | CT 17-36 |
| 43 | Humasis Co., Ltd. | Humasis COVID-19 Ag Test | 88,2% | 21,7% | 0,0% | 40,0% |
| 44 | IVC Pragen Healthcare | GenBody COVID-19 Ag | 94,4% | 26,1% | 0,0% | 46,0% |
| 45 | Jiangsu Diagnostics Biotechnology Co., Ltd | COVID-19 Antigen Rapid Test Cassette (Colloidal Gold) | 100,0% | 78,3% | 0,0% | 68,0% |
| 46 | Jiangsu Medomics Medical Technology Co., Ltd | SARS-CoV-2-Antigen-Testkit (LFIA) | 94,1% | 21,7% | 0,0% | 42,0% |
| 47 | Joinstar Biomedical Technology Co., Ltd (CIV care impuls Vertrieb) | COVID-19 Antigen Schnelltest (Colloidal Gold) | 100,0% | 60,9% | 0,0% | 64,0% |
| 48 | Labnovation Technologies, Inc. | Labnovation SARS-CoV-2 Antigen Rapid Test Kit (Immunochromatography) | 94,1% | 17,4% | 0,0% | 40,0% |
| 49 | Lumigenex (Suzhou) Co., Ltd. | PocRoc SARS-CoV-2, Antigen Schnelltest Set (Kolloidales Gold) | 100,0% | 65,2% | 0,0% | 64,0% |
| 50 | LumiQuick Diagnostics, Inc. | QuickProfile Covid-19 Antigen Test Card | 100,0% | 91,3% | 20,0% | 80,0% |
| 51 | LumiraDX | LumiraDx SARS-CoV-2 Ag Test | 100,0% | 52,2% | 0,0% | 60,0% |
| 52 | MEDsan GmbH | MEDsan® SARS-CoV-2 Antigen Rapid Test | 100,0% | 47,8% | 0,0% | 58,0% |
| 53 | Merlin Biomedical (Xiamen) Co., Ltd. | SARS-CoV-2 Antigen Rapid Test Cassette | 100,0% | 82,6% | 0,0% | 72,0% |
| 54 | Mölab GmbH | mö-screen Corona Antigen Test | 100,0% | 47,8% | 0,0% | 58,0% |
| 55 | MP Biomedicals Germany GmbH | Rapid SARS-CoV-2 Antigen Test Card | 100,0% | 43,5% | 0,0% | 54,0% |
| 56 | nal von minden gmbh | NADAL® COVID-19 Ag Schnelltest | 83,3% | 13,0% | 0,0% | 36,0% |
| 57 | Nanjing Norman Biological Technology Co.,Ltd | Novel Coronavirus (2019-nCoV) Antigen Testing Kit (Colloidal Gold) | 94,1% | 26,1% | 0,0% | 44,0% |
| 58 | NanoEntek Inc | FREND™ COVID-19 Ag | 88,2% | 8,7% | 0,0% | 34,0% |
| 59 | Nantong Diagnos Biotechnology Co., Ltd. | COVID-19 Antigen Saliva Test Kit (Colloidal Gold) | 100,0% | 56,5% | 0,0% | 60,0% |
| 60 | New Gene (Hangzhou) Bioengineering Co., Ltd. | Covid-19-Antigen-Testkit | 100,0% | 87,0% | 20,0% | 78,0% |
| 61 | Novatech Tibbi Cihaz Ürünleri San. Ve Tic.A.S. | novacheck®-Ag SARS-CoV-2 Covid-19 Antigen Rapid Test | 94,1% | 21,7% | 0,0% | 42,0% |
| 62 | Oncosem Onkolojik Sistemler San. Ve Tic. A.S. | CAT Antigen Covid Rapid Test | 94,1% | 30,4% | 0,0% | 46,0% |
| 63 | PCL, Inc. | PCL COVID19 Ag Gold Saliva | 100,0% | 52,2% | 0,0% | 58,0% |
| 64 | PerGrande BioTech Development Co., Ltd. | SARS-CoV-2 Antigen Detection Kit (Colloidal Gold Immunochromatographic Assay) | 100,0% | 17,4% | 0,0% | 42,0% |
| 65 | Precision Biosensor Inc. (Axon Lab AG) | Exdia COVID-19-Ag-Test | 100,0% | 60,9% | 0,0% | 64,0% |
| 66 | ProGnosis Biotech | Rapid Test Ag 2019-nCoV | 94,1% | 65,2% | 10,0% | 64,0% |
| 67 | Quidel Corporation | Sofia SARS Antigen FIA | 88,9% | 8,7% | 0,0% | 36,0% |
| 68 | Qingdao Hightop Biotech Co., Ltd. | Hightop SARS-CoV-2 (Covid-19) Antigen Rapid Test | 100,0% | 43,5% | 0,0% | 54,0% |
| 69 | R-Biopharm AG | RIDA®QUICK SARS-CoV-2 Antigen | 100,0% | 17,4% | 0,0% | 44,0% |
| 70 | Safecare Biotech Hangzhou Co., Ltd. | Safecare COVID-19 Ag Rapid Test Kit (Swab) | 100,0% | 60,9% | 0,0% | 62,0% |
| 71 | Salofa OY | salocor SARS-CoV-2 Antigen Rapid Test Cassette (Nasopharyngeal swab) | 82,4% | 13,0% | 0,0% | 34,0% |
| 72 | ScheBo Biotech AG | ScheBo SARS-CoV-2 Quick Antigen | 100,0% | 91,3% | 10,0% | 78,0% |
| 73 | SD BIOSENSOR (Roche Diagnostics GmbH) | SARS-CoV-2 Rapid Antigen Test | 88,9% | 30,4% | 0,0% | 46,0% |
| 74 | SD BIOSENSOR | STANDARD™ Q COVID-19 Ag Test | 88,9% | 30,4% | 0,0% | 46,0% |
| 75 | SD BIOSENSOR | STANDARD™ F COVID-19 Ag FIA | 100,0% | 65,2% | 0,0% | 66,0% |
| 76 | SGA Mühendislik DAN. EG. İcve DIS.Ltd.STI | V-Chek SARS-CoV-2 Rapid Ag Test Kit (Colloidal Gold) | 94,1% | 26,1% | 0,0% | 44,0% |
| 77 | Shenzhen Lvshiyuan Biotechnology Co., Ltd. | Green Spring SARS-CoV-2 Antigen Rapid Test Kit (Colloidal Gold) | 100,0% | 95,7% | 40,0% | 86,0% |
| 78 | Shenzhen Microprofit Biotech Co., Ltd | fluorecare COVID-19 SARS-CoV-2 Spike Protein Test Kit (Colloidal Gold Chromatographic Immunoassay) | 100,0% | 47,8% | 10,0% | 58,0% |
| 79 | Shenzhen Watmind Medical Co.,Ltd. | SARS-CoV-2 Ag Diagnostic Test Kit (Colloidal Gold) | 100,0% | 95,7% | 20,0% | 82,0% |
| 80 | Shenzhen Watmind Medical Co.,Ltd. | SARS-CoV-2 Ag Diagnostic Test Kit (Immuno-fluorescence) | 100,0% | 60,9% | 0,0% | 62,0% |
| 81 | Shenzhen Zhenrui Biotech co.Ltd. | Zhenrui COVID-19 (SARS-COV-2) Antigen Test Kits | 82,4% | 13,0% | 0,0% | 34,0% |
| 82 | Siemens Healthineers | CLINITEST® Rapid COVID-19 Antigen Test | 100,0% | 87,0% | 0,0% | 76,0% |
| 83 | Sugentech, Inc. | SGTi-flex COVID-19 Ag | 100,0% | 73,9% | 0,0% | 68,0% |
| 84 | Toda Pharma | Toda Coronadiag Ag | 100,0% | 95,7% | 40,0% | 86,0% |
| 85 | Triplex International Biosciences (China) Co., Ltd. | SARS-CoV-2 Antigen Rapid Test Kit | 100,0% | 87,0% | 20,0% | 78,0% |
| 86 | ulti med Products (Deutschland) GmbH | COVID-19 Antigen Speicheltest (Immunochromatographie) | 100,0% | 95,7% | 20,0% | 82,0% |
| 87 | Vitrosens Biyoteknoloji Ltd. Sti | RapidFor SARS-CoV-2 Rapid Antigen Test Colloidal Gold | 100,0% | 30,4% | 0,0% | 48,0% |
|  |  |  | CT <25 | CT 25-30 | CT >30 | CT 17-36 |
| 88 | Wantai (Beijing Wantai Biological Pharmacy Enterprise Co., Ltd.) | SARS-CoV-2 Ag Rapid Test (FIA) | 100,0% | 78,3% | 0,0% | 72,0% |
| 89 | Wuhan EasyDiagnosis Biomedicine Co., Ltd | COVID-19 (SARS-CoV-2) Antigen Test Kit | 100,0% | 73,9% | 0,0% | 68,0% |
| 90 | Wuhan Life Origin Biotech Joint Stock Co., Ltd. | SARS-CoV-2 Antigen Assay Kit (Immunochromatography) | 100,0% | 56,5% | 0,0% | 60,0% |
| 91 | Wuhan UNscience Biotechnology Co., Ltd. | SARS-CoV-2 Antigen Rapid Test Kit | 88,2% | 17,4% | 0,0% | 38,0% |
| 92 | Xiamen Boson Biotech Co., Ltd | SARS-CoV-2 Antigen Schnelltest | 100,0% | 43,5% | 0,0% | 54,0% |
| 93 | Xiamen WIZ Biotech Co., Ltd. | Wizbiotech SARS-CoV-2 Antigen Rapid Test | 88,2% | 13,0% | 0,0% | 36,0% |
| 94 | Zet Medikal Tekstil Dis Ticaret Ltd. STI. | softec SARS COV-2 (Covid-19) Antigen Test Kit | 82,4% | 21,7% | 10,0% | 40,0% |
| 95 | Zhejiang Anji Saianfu Biotech Co.,Ltd. | reOpenTest COVID-19 Antigen Rapid Test (Colloidal Gold) | 94,1% | 30,4% | 0,0% | 46,0% |
| 96 | Zhejiang Orient Gene Biotech Co.,Ltd | Coronavirus Ag Rapid Test Cassette (Swab) | 100,0% | 87,0% | 0,0% | 76,0% |

**Table 2.** Comparative evaluation results of SARS-CoV-2 antigen RDT missing the sensitivity criteria. (in alphabetical order of manufacturers)

| No. | Manufacturer | Test name | Sensitivity |  |  |  |
| --- | --- | --- | --- | --- | --- | --- |
|  |  |  | CT <25 | CT 25-30 | CT >30 | CT 17-36 |
| 1 | Acro Biotech Inc | Acro COVID-19 Antigen Rapid Test | 16,7% | 0,0% | 0,0% | 6,0% |
| 2 | Aikang Diagnostics Co., Ltd. | SARS-CoV-2 Antigen Test Kit (Immunochromatography) | 11,8% | 0,0% | 0,0% | 4,0% |
| 3 | Beijing Savant Biotechnology Co., Ltd | New Coronavirus (SARS-CoV-2) N Protein Detection Kit (Fluorescence Immunchromatography) | 0,0% | 0,0% | 0,0% | 0,0% |
| 4 | Certest Biotec S. L. | CerTest SARS-CoV-2 | 29,4% | 0,0% | 0,0% | 10,0% |
| 5 | Coris Bioconcept | COVID-19 Ag Respi-Strip | 33,3% | 0,0% | 0,0% | 12,0% |
| 6 | Hangzhou AllTest Biotech Co. Ltd. | COVID-19 AG AllTest | 16,7% | 0,0% | 0,0% | 6,0% |
| 7 | Hangzhou Biotest Biotech Co., Ltd. | Lumiratek SARS-CoV-2 Antigen Rapid Test Cassette | 29,4% | 0,0% | 0,0% | 10,0% |
| 8 | Hangzhou Genesis Biocontrol Co., Ltd | KaiBiLi COVID-19 Antigen Rapid Test Device | 52,9% | 0,0% | 0,0% | 18,0% |
| 9 | Hangzhou Realy Tech Co., Ltd. | Novel Coronavirus (SARS-Cov-2) Antigen Rapid Test Cassette (swab) | 58,8% | 0,0% | 0,0% | 20,0% |
| 10 | Inzek International Trading | Biozek medical COVID-19 Antigen Rapid Test Cassette | 52,9% | 0,0% | 0,0% | 18,0% |
| 11 | Joinstar Biomedical Technology Co., Ltd | COVID-19 Antigen Rapid Test (Latex) | 0,0% | 0,0% | 0,0% | 0,0% |
| 12 | Joysbio (Tianjin) Biotechnology Co., Ltd. | Joysbio SARS-CoV-2 Antigen Rapid Test Kit (Colloidal Gold) | 47,1% | 4,3% | 0,0% | 18,0% |
| 13 | Lionex GmbH | Lionex COVID-19 Ag Rapid Test | 0,0% | 0,0% | 0,0% | 0,0% |
| 14 | Medicon Co., Ltd. | Trueline COVID-19 Ag Rapid Test | 58,8% | 4,3% | 0,0% | 22,0% |
| 15 | Mexacare GmbH Heidelberg | QuickTestCorona COVID-19 Antigen Schnelltest | 52,9% | 4,3% | 0,0% | 20,0% |
| 16 | nal von minden GmbH | dedicio Medical Test COVID-19 Ag plus Test | 35,3% | 0,0% | 0,0% | 12,0% |
| 17 | Rapigen | Biocredit COVID-19 Ag | 16,7% | 0,0% | 0,0% | 6,0% |
| 18 | Servoprax | Cleartest Coronaantigen | 66,7% | 0,0% | 0,0% | 24,0% |
| 19 | Spring Healthcare Services SP zoo | SARS-Cov-2 Antigen Rapid Test Cassette (swab) | 29,4% | 0,0% | 0,0% | 10,0% |
| 20 | SureScreen Diagnostics Ltd | COVID-19 Antigen Rapid Test Cassette | 52,9% | 0,0% | 0,0% | 18,0% |
| 21 | TaiDoc TechnologyCorp. | FORA COVID-19 ANTIGEN RAPID TEST | 27,8% | 0,0% | 0,0% | 10,0% |
| 22 | Unioninvest | Unibioscience COVID-19 Rapid Antigen Test | 0,0% | 0,0% | 0,0% | 0,0% |
| 23 | VivaChek Biotech (Hangzhou) Co.Ltd. | VivaDiag SARS-CoV-2 Ag Rapid Test | 50,0% | 0,0% | 0,0% | 18,0% |
| 24 | VivaChek Biotech (Hangzhou) Co.Ltd. | VivaDiag Pro SARS-CoV-2 Ag Rapid Test | 64,7% | 0,0% | 0,0% | 22,0% |
| 25 | W.H.P.M, Inc | First SIGN SARS-CoV-2 Antigen Test | 47,1% | 0,0% | 0,0% | 16,0% |
| 26 | Xiamen Zhongsheng Langjie Biotechnology Co., Ltd | Covid-19 Antigen Test Cassette | 11,8% | 0,0% | 0,0% | 4,0% |

The 96 tests meeting the sensitivity criteria were reactive with between 14 and 41 members of the 50 members panel (supplemental Table 1). On average throughout all successful tests, 27 panel members were reactive. Overall reactivity of SARS-CoV-2 Ag RDT strongly followed the analyte concentration throughout the panel, justifying the conclusions of this study (supplemental Table 1).

The 26 SARS-CoV-2 Ag RDT missing the sensitivity criteria either failed completely (2 tests with 0 reactives) or were reactive with 2 to 12 (average 6.3) panel members. Again, reactivity was dependent on the analyte concentration throughout the panel members (supplemental Table 2). Two tests failed because of constant faint background reactivity throughout all panel members; this background reactivity was also seen when using pure extraction buffer and was thus not caused by the panel composition (data not shown).

## Discussion

There is convincing evidence that infectivity of SARS-CoV-2 correlates directly with high viral loads in respiratory specimens of acutely infected persons. It has therefore been suggested to use antigen tests rather to detect potential infectivity to help to control the spread of infection than for the purpose of clinical diagnosis. Ag RDTs are designed to detect SARS-CoV-2 antigens in respective samples and have become a key part of testing strategies in many countries since fall 2020. Hundreds of different Ag RDTs, most often of East-Asian origin, are available on the Common Market in Europe. Nearly all tests state in their IFUs sensitivity values of >90% for PCR-confirmed specimens. Such statements are unreliable and in strong contrast to the results of our study and to independent evaluations. Their only explanation is a strong preselection of high-positive specimens.

Lack of independent evaluation combined with unreliable statements of quality features led the German Ministry of Health to request a comparative evaluation of the sensitivity of test kits offered in Germany. At the time being there are no EU-wide requirements for quality features of COVID-19 IVDs such as a defined minimum sensitivity or minimum specificity, and manufacturers may themselves certify their devices to comply with basic requirements of the IVDD. Therefore, it is mainly left to individual Member States or international organizations to define minimum requirements for acceptance of respective tests.

In Germany, the Ministry of Health decided to link the reimbursement of SARS-CoV-2 Ag RDT to quality requirements to be fulfilled by acceptable devices. Minimum requirements were jointly formulated by Paul-Ehrlich-Institute and Robert Koch-Institute and state for SARS-CoV-2 Ag RDTs a minimum sensitivity of 80% for PCR positive specimens obtained within the first seven days after symptom onset; the minimum specificity was defined as >97%, and for both requirements a study population of at least 100 persons is required. Analogous requirements of SARS-CoV-2 Ag RDTs have been proposed by the World Health Organization for the emergency use listing (EUL) (6), the US Food and Drug Administration (FDA) (7), the European Center for Disease Control (ECDC) (8), the Swiss Authority Bundesamt für Gesundheit (BAG) (9) or the non-governmental Foundation for Innovative New Diagnostics (FIND) (10). With mandatory application of the EU IVD Regulation (IVDR) for SARS-CoV-2 Ag RDT their regulatory oversight will improve in near future. In this context it might be of interest that so-called Common Specifications have already been drafted for SARS-CoV-2 IVD. Although these draft quality requirements are still under consultation and not yet in effect, they will become essential prerequisites for future CE-marking of respective devices.

For the time being, successful participation in a comparative evaluation of the crucial test feature sensitivity was installed as another precondition for reimbursement of SARS-CoV-2 Ag RDT by the German healthcare system. The first round of this evaluation is summarized in this manuscript.

The sensitivity requirement defined for a successful outcome in the comparative evaluation study (>75% sensitivity for CT<25) is in line with international requirements mentioned above, as far as clinical specimens from acutely infected individuals are comparable to the members of our evaluation panel. We followed routine use of the tests as far as possible, including pre-analytical steps like antigen absorption of the test-specific swabs, and subsequent elution into the test-specific buffer. The vast majority of Ag RDTs included in our study showed sufficient sensitivity according to our criteria. Nevertheless, the results showed a wide range of varying sensitivities. There were few tests with fairly high and many tests with sufficient sensitivity, but also quite a few tests, which did not meet the minimum criterion. The study shows that the majority of SARS-CoV-2 Ag RDTs correctly identify high viral loads of <CT 25 (>10^6^ virus RNA copies/mL) in samples from the respiratory tract with a sensitivity of >75%, supporting their use in the early symptomatic phase. However, although sensitivity often declined with CT>25, there were also few SARS-CoV-2 Ag RDTs with quite high sensitivity, even 100% for CT<30, or up to 86% for the complete CT range (CT 17-36). There are scientific publications of further independent head-to-head evaluations for SARS-CoV-2 Ag RDT which, however, are limited to the comparison of only few tests (11-17). Respective conclusions based on clinical specimens are consistent with our results, and the sensitivity ranking of different tests was widely in line with our results based on the evaluation panel.

Since most of the SARS-CoV-2 Ag RDTs offered in Europe are provided without a readout device, visual interpretation of test results is indispensable. We would like to emphasize that few discrepant tests results obtained by two experienced lab technicians were reported. These equivocal results were ultimately interpreted as reactive, in favor of the tests under investigation. However, visual readout and subjective interpretation of faint test lines, potentially caused by borderline concentration of the analyte, presents a challenge for less experienced users, e.g. lay persons using Ag RDTs as self tests.

A limitation of the study is its spot check nature since it cannot address variations between different batches of the same product, or variations between different test locations (see also the tandem publication of Puyskens et al).

In conclusion, by using the same panel for a large number of different SARS-CoV-2 Ag RDT we were able to evaluate the comparative performance of the different tests under the same conditions. The evaluation panel proved to be accurate for sensitivity differentiation of SARS-CoV-2 Ag RDTs, distinguishing better performing from less suitable tests. The continuation of the comparative evaluation is needed cope with the rapidly growing market of SARS-CoV-2 Ag RDT. Since the panel has now been exhausted, we will continue the evaluation with a new set of samples with similar features, accurately calibrated against its predecessor.

## Data Availability

All relevant data are contained in the manuscript

