## Supplemental Table 1 for "Comparative sensitivity evaluation for 122 CE-marked SARS-CoV-2 antigen rapid tests"

Comparative evaluation results of SARS-CoV-2 antigen RDT passing the sensitivity criteria

(in alphabetical order of manufacturers)

| | | | | 1 | 2 | 3 | 4 | 5 | 6 | 7 | 8 | 9 | 10 | 11 | 12 | 13 | 14 | 15 | 16 | 17 |
|---|---|---|---|---|---|---|---|---|---|---|---|---|---|---|---|---|---|---|---|---|
| | | **Manufacturer** | | Abbott Rapid Diagnostics Jena GmbH | ACON Biotech (Hangzhou) Co., Ltd | Aesku Diagnostics GmbH | Affimedix | Amazing Biotech (Shanghai) Co., Ltd | Ameda Labordiagnostik GmbH | AmonMed (Xiamen) Biotechnology Co.,Ltd. | Anbio (Xiamen) Biotechnology Co., Ltd | Anhui Deepblue Medical Technology Co., Ltd. | ASAN PHARM.CO.,LTD. | Atlas Link Technology Co.,Ltd. | Avalun | AXIOM Gesellschaft für Diagnostica und Biochemica mbH | Azure Biotech Inc. | Becton Dickinson | Beijing Beier Bioengineering Co., Ltd. | Beijing Hotgen Biotech Co., Ltd. |
| | | **Test name** | | Panbio™ Covid-19 Ag Rapid Test Device | Flowflex SARS-CoV-2-Antigenschnelltest (Nasopharynxtupfer) | Aesku Rapid SARS-CoV-2 Rapid Test | TestNOW® - COVID-19 Antigen | CoroVisio Covid-19 Ag Versieglungsröhrchen Teststreifen (Kolloidales Gold) | AMP Rapid Test SARS-CoV-2 Ag | COVID-19 Antigen Rapid Test Kit (Colloidal Gold) | Rapid Covid-19 Antigen Test (Colloidal Gold ) | COVID-19 (SARS CoV-2) Antigen Test Kit (Colloidal Gold) | Asan Easy Test COVID-19 Ag | Nova Test SARS-CoV-2 Antigen Rapid Test Kit | Ksmart® SARS-COV2 Antigen Rapid Test | Axiom Diagnostics COVID-19 Ag Schnelltest | Dia Sure Covid-19 Antigen Rapid Test Device (Nasopharyngeal/ Oropharyngeal Swab) | BD Veritor™ System for Rapid Detection of SARS-CoV-2 | Covid-19 Antigen Schnelltest | Neuartiges Coronavirus (2019-nCoV)-Antigentest (Kolloidales Gold) |
| **Panel 1 V1** | | **Panel 1 V2** | | Panel 1 V1 | Panel 1 V2 | Panel 1 V2 | Panel 1 V1 | Panel 1 V2 | Panel 1 V2 | Panel 1 V2 | Panel 1 V2 | Panel 1 V2 | Panel 1 V2 | Panel 1 V2 | Panel 1 V2 | Panel 1 V2 | Panel 1 V2 | Panel 1 V1 | Panel 1 V1 | Panel 1 V2 |
| **Pool Nr.** | **CT** | **Pool Nr.** | **CT** | | | | | | | | | | | | | | | | | |
| 1 | 17,55 | 1 | 17,31 | 1 | 1 | 1 | 1 | 1 | 1 | 1 | 1 | 1 | 1 | 1 | 1 | 1 | 1 | 1 | 1 | 1 |
| 6 | 20,20 | 2 | 19,08 | 1 | 1 | 1 | 1 | 1 | 1 | 1 | 1 | 1 | 1 | 1 | 1 | 1 | 1 | 1 | 1 | 1 |
| 5 | 20,28 | 3 | 19,62 | 1 | 1 | 1 | 1 | 1 | 1 | 1 | 1 | 1 | 1 | 1 | 1 | 1 | 1 | 1 | 1 | 1 |
| 3 | 20,38 | 5 | 20,60 | 1 | 1 | 1 | 1 | 1 | 1 | 1 | 1 | 1 | 1 | 1 | 1 | 1 | 1 | 1 | 1 | 1 |
| 2 | 20,54 | 4 | 20,61 | 1 | 1 | 1 | 1 | 1 | 1 | 1 | 1 | 1 | 1 | 1 | 1 | 1 | 1 | 1 | 1 | 1 |
| 4 | 20,98 | 6 | 21,21 | 1 | 1 | 1 | 1 | 1 | 1 | 1 | 1 | 1 | 1 | 1 | 1 | 1 | 1 | 1 | 1 | 1 |
| 7 | 21,71 | 12 | 22,12 | 1 | 1 | 1 | 1 | 0 | 1 | 1 | 1 | 1 | 1 | 1 | 1 | 1 | 1 | 1 | 1 | 1 |
| 12 | 21,82 | 7 | 22,15 | 1 | 1 | 1 | 1 | 1 | 1 | 1 | 1 | 1 | 1 | 1 | 1 | 1 | 1 | 1 | 1 | 1 |
| 8 | 21,95 | 8 | 22,32 | 1 | 1 | 1 | 1 | 0 | 1 | 1 | 1 | 1 | 1 | 1 | 1 | 1 | 1 | 1 | 1 | 1 |
| 9 | 22,14 | 16 | 22,88 | 1 | 1 | 1 | 1 | 1 | 1 | 1 | 1 | 1 | 1 | 1 | 1 | 1 | 1 | 1 | 1 | 1 |
| 11 | 22,34 | 9 | 23,13 | 1 | 1 | 1 | 1 | 1 | 1 | 1 | 1 | 1 | 1 | 1 | 1 | 1 | 1 | 0 | 1 | 1 |
| 16 | 22,55 | 11 | 23,13 | 1 | 1 | 0 | 1 | 1 | 1 | 1 | 1 | 1 | 1 | 1 | 0 | 1 | 1 | 1 | 1 | 1 |
| 10 | 22,88 | 10 | 23,21 | 1 | 1 | 1 | 1 | 1 | 1 | 1 | 1 | 1 | 1 | 1 | 1 | 1 | 1 | 1 | 0 | 1 |
| 13 | 23,32 | 15 | 24,38 | 1 | 1 | 0 | 1 | 0 | 1 | 1 | 1 | 1 | 1 | 1 | 1 | 1 | 0 | 1 | 1 | 1 |
| 17 | 24,00 | 23 | 24,45 | 1 | 1 | 0 | 1 | 0 | 1 | 1 | 1 | 1 | 1 | 1 | 1 | 1 | 0 | 1 | 1 | 1 |
| 23 | 24,04 | 17 | 24,81 | 1 | 0 | 1 | 1 | 1 | 1 | 1 | 1 | 1 | 1 | 1 | 1 | 1 | 0 | 0 | 0 | 1 |
| 15 | 24,14 | 14 | 24,97 | 1 | 1 | 1 | 1 | 1 | 1 | 1 | 1 | 1 | 1 | 1 | 1 | 1 | 0 | 0 | 0 | 1 |
| 14 | 24,28 | 25 | 25,07 | 1 | 0 | 0 | 1 | 0 | 1 | 1 | 1 | 1 | 1 | 1 | 1 | 1 | 0 | 1 | 0 | 1 |
| 27 | 25,14 | 24 | 25,20 | 1 | 0 | 0 | 1 | 1 | 1 | 1 | 1 | 1 | 1 | 1 | 0 | 1 | 0 | 0 | 0 | 1 |
| 24 | 25,24 | 13 | 25,29 | 1 | 1 | 1 | 1 | 1 | 1 | 1 | 1 | 1 | 1 | 1 | 1 | 1 | 1 | 0 | 0 | 1 |
| 31 | 25,27 | 19 | 25,45 | 0 | 0 | 1 | 0 | 0 | 1 | 1 | 1 | 1 | 1 | 1 | 0 | 1 | 0 | 0 | 0 | 1 |
| 18 | 25,30 | 21 | 25,95 | 1 | 0 | 0 | 1 | 0 | 1 | 1 | 0 | 1 | 1 | 1 | 0 | 0 | 0 | 1 | 0 | 1 |
| 26 | 25,47 | 27 | 26,12 | 1 | 0 | 0 | 1 | 0 | 1 | 1 | 1 | 1 | 1 | 1 | 0 | 1 | 0 | 1 | 0 | 1 |
| 19 | 25,50 | 31 | 26,24 | 1 | 0 | 0 | 1 | 0 | 1 | 1 | 1 | 0 | 1 | 1 | 0 | 0 | 0 | 0 | 0 | 0 |
| 21 | 25,54 | 26 | 26,32 | 1 | 0 | 0 | 1 | 0 | 1 | 1 | 0 | 0 | 1 | 0 | 0 | 1 | 0 | 0 | 0 | 0 |
| 22 | 25,87 | 32 | 26,64 | 1 | 0 | 0 | 1 | 0 | 1 | 1 | 0 | 0 | 0 | 0 | 0 | 0 | 0 | 0 | 0 | 0 |
| 20 | 26,27 | 35 | 26,66 | 1 | 0 | 0 | 1 | 0 | 1 | 1 | 1 | 0 | 1 | 1 | 0 | 0 | 1 | 0 | 0 | 1 |
| 32 | 26,44 | 36 | 27,05 | 0 | 0 | 0 | 0 | 0 | 1 | 1 | 1 | 0 | 1 | 1 | 0 | 0 | 1 | 0 | 0 | 0 |
| 35 | 27,04 | 30 | 27,24 | 1 | 0 | 0 | 1 | 0 | 1 | 1 | 1 | 0 | 1 | 0 | 0 | 1 | 0 | 0 | 0 | 1 |
| 28 | 27,14 | 29 | 27,34 | 0 | 0 | 0 | 0 | 0 | 1 | 1 | 0 | 0 | 1 | 1 | 0 | 1 | 0 | 0 | 0 | 0 |
| 29 | 27,15 | 28 | 27,41 | 1 | 0 | 0 | 0 | 0 | 1 | 1 | 0 | 0 | 0 | 0 | 0 | 1 | 0 | 0 | 0 | 0 |
| 40 | 27,65 | 22 | 27,42 | 0 | 0 | 1 | 0 | 0 | 1 | 1 | 1 | 1 | 1 | 1 | 0 | 1 | 0 | 0 | 0 | 1 |
| 34 | 27,89 | 34 | 27,82 | 1 | 0 | 0 | 0 | 0 | 0 | 0 | 0 | 0 | 0 | 0 | 0 | 0 | 0 | 0 | 0 | 0 |
| 36 | 28,13 | 40 | 28,19 | 1 | 0 | 0 | 1 | 0 | 0 | 0 | 0 | 0 | 0 | 0 | 0 | 0 | 0 | 0 | 0 | 0 |
| 38 | 28,14 | 18 | 28,33 | 1 | 0 | 1 | 0 | 0 | 1 | 1 | 1 | 1 | 1 | 1 | 1 | 1 | 0 | 0 | 0 | 1 |
| 42 | 28,43 | 33 | 28,92 | 0 | 0 | 0 | 0 | 0 | 0 | 1 | 0 | 0 | 0 | 0 | 0 | 0 | 0 | 0 | 0 | 0 |
| 30 | 28,86 | 38 | 29,36 | 1 | 0 | 0 | 0 | 0 | 1 | 1 | 0 | 0 | 1 | 1 | 0 | 0 | 0 | 0 | 0 | 0 |
| 33 | 28,96 | 20 | 29,46 | 0 | 0 | 0 | 0 | 0 | 1 | 1 | 1 | 1 | 1 | 1 | 0 | 1 | 0 | 0 | 0 | 1 |
| 44 | 29,24 | 42 | 29,48 | 0 | 0 | 0 | 0 | 0 | 0 | 1 | 0 | 0 | 0 | 0 | 0 | 0 | 0 | 0 | 0 | 0 |
| 25 | 29,70 | 44 | 29,51 | 0 | 0 | 0 | 1 | 0 | 0 | 0 | 0 | 0 | 0 | 0 | 0 | 0 | 0 | 0 | 0 | 0 |
| 39 | 29,76 | 39 | 30,12 | 0 | 0 | 0 | 0 | 0 | 0 | 1 | 0 | 0 | 0 | 0 | 0 | 0 | 0 | 0 | 0 | 0 |
| 45 | 30,10 | 37 | 30,13 | 0 | 0 | 0 | 0 | 0 | 0 | 1 | 0 | 0 | 0 | 0 | 0 | 0 | 0 | 0 | 0 | 0 |
| 41 | 30,13 | 41 | 30,14 | 0 | 0 | 0 | 0 | 0 | 0 | 0 | 0 | 0 | 0 | 0 | 0 | 0 | 0 | 0 | 0 | 0 |
| 37 | 30,54 | 45 | 31,19 | 0 | 0 | 0 | 0 | 0 | 0 | 0 | 0 | 0 | 0 | 0 | 0 | 0 | 0 | 0 | 0 | 0 |
| 43 | 31,05 | 48 | 31,19 | 0 | 0 | 0 | 0 | 0 | 0 | 0 | 0 | 0 | 0 | 0 | 0 | 0 | 0 | 0 | 0 | 0 |
| 46 | 31,54 | 46 | 31,34 | 0 | 0 | 0 | 0 | 0 | 0 | 1 | 0 | 0 | 0 | 0 | 0 | 0 | 1 | 0 | 0 | 0 |
| 48 | 32,06 | 43 | 31,61 | 0 | 0 | 0 | 0 | 0 | 0 | 0 | 0 | 0 | 0 | 0 | 0 | 0 | 1 | 0 | 0 | 0 |
| 47 | 35,19 | 47 | 34,55 | 0 | 0 | 0 | 0 | 0 | 0 | 0 | 0 | 0 | 0 | 0 | 0 | 0 | 0 | 0 | 0 | 0 |
| 49 | 35,22 | 50 | 35,83 | 0 | 0 | 0 | 0 | 0 | 0 | 0 | 0 | 0 | 0 | 0 | 0 | 0 | 0 | 0 | 0 | 0 |
| 50 | 36,36 | 49 | 36,04 | 0 | 0 | 0 | 0 | 0 | 0 | 0 | 0 | 0 | 0 | 0 | 0 | 0 | 0 | 0 | 0 | 0 |

Supplemental Table 1

Comparative evaluation results of SARS-CoV-2 antigen RDT passing the sensitivity criteria

(in alphabetical order of manufacturers)

| | | | | 18 | 19 | 20 | 21 | 22 | 23 | 24 | 25 | 26 | 27 | 28 | 29 | 30 | 31 | 32 | 33 | 34 |
|---|---|---|---|---|---|---|---|---|---|---|---|---|---|---|---|---|---|---|---|---|
| | | **Manufacturer** | | Beijing Lepu Medical Technology Co., Ltd | Beijing Tigsun Diagnostics Co.;Ltd. | BIOMERICA Inc. | BIONOTE | BioRepair GmbH | BIOSYNEX SWISS SA | BTNX, Inc. (Biotrend Chemikalien GmbH) | Chil Tibbi Mal. San. Tic. Ltd. Şti | Core Technology Co., Ltd. | DNA Diagnostic A/S. | Edinburgh Genetics Limited | Eurobio Scientific | Fujirebio Inc. (Mast Diagnostica GmbH) | Genrui Biotech Inc. | GenSure Biotech Inc. | Getein Biotech, Inc. | Green Cross Medical Science Corp. (Weko Pharma GmbH) |
| | | **Test name** | | SARS-CoV-2 Antigen Rapid Test Kit | Tigsun COVID-19 Saliva Antigen Rapid Test | COVID-19-Antigen-Schnelltest (Nasopharyngeal-Abstrich) | NowCheck® COVID-19 Ag Test | Covid 19 Antigen Schnelltest | BIOSYNEX COVID-19 Ag BSS | Rapid Response COVID-19 Rapid Test Device | COVID-19 Antigen Schnell Test (Nasopharyngeal / Oropharyngeal Tupfer - Kassette) | Canea COVID-19 Antigen Schnelltest | Covid-19 Antigen Detection Kit | Edinburgh Genetics ActivXpress+ COVID-19 Antigen Complete Testing Kit | EBS SARS-CoV-2 Ag Rapid Test | ESPLINE® SARS-CoV-2 | Genrui SARS-CoV-2 Antigen Test Kit (Colloidal Gold) | GensureTM COVID-19 Antigen Rapid Test Kit | One Step Test for SARS-CoV-2 Antigen (Colloidal Gold) | Genedia W Covid-19 Ag |
| **Panel 1 V1** | | **Panel 1 V2** | | Panel 1 V2 | Panel 1 V2 | Panel 1 V2 | Panel 1 V1 | Panel 1 V2 | Panel 1 V1 | Panel 1 V2 | Panel 1 V2 | Panel 1 V2 | Panel 1 V2 | Panel 1 V2 | Panel 1 V2 | Panel 1 V1 | Panel 1 V2 | Panel 1 V2 | Panel 1 V2 | Panel 1 V1 |
| **Pool Nr.** | **CT** | **Pool Nr.** | **CT** | | | | | | | | | | | | | | | | | |
| 1 | 17,55 | 1 | 17,31 | 1 | 1 | 1 | 1 | 1 | 1 | 1 | 1 | 1 | 1 | 1 | 1 | 1 | 1 | 1 | 1 | 1 |
| 6 | 20,20 | 2 | 19,08 | 1 | 1 | 1 | 1 | 1 | 1 | 1 | 1 | 1 | 1 | 1 | 1 | 1 | 1 | 1 | 1 | 1 |
| 5 | 20,28 | 3 | 19,62 | 1 | 1 | 1 | 1 | 1 | 1 | 1 | 1 | 1 | 1 | 1 | 1 | 1 | 1 | 1 | 1 | 1 |
| 3 | 20,38 | 5 | 20,60 | 1 | 1 | 1 | 1 | 1 | 1 | 1 | 1 | 1 | 1 | 1 | 1 | 1 | 1 | 1 | 1 | 1 |
| 2 | 20,54 | 4 | 20,61 | 1 | 1 | 1 | 1 | 1 | 1 | 1 | 1 | 1 | 1 | 1 | 1 | 1 | 1 | 1 | 1 | 1 |
| 4 | 20,98 | 6 | 21,21 | 1 | 1 | 1 | 1 | 1 | 1 | 1 | 1 | 1 | 1 | 1 | 1 | 1 | 1 | 1 | 1 | 1 |
| 7 | 21,71 | 12 | 22,12 | 1 | 1 | 1 | 1 | 1 | 1 | 1 | 1 | 1 | 1 | 1 | 1 | 1 | 1 | 1 | 1 | 1 |
| 12 | 21,82 | 7 | 22,15 | 1 | 1 | 1 | 1 | 1 | 1 | 1 | 1 | 1 | 1 | 1 | 1 | 1 | 1 | 1 | 1 | 1 |
| 8 | 21,95 | 8 | 22,32 | 1 | 1 | 1 | 1 | 1 | 1 | 1 | 1 | 1 | 1 | 1 | 1 | 1 | 1 | 1 | 1 | 1 |
| 9 | 22,14 | 16 | 22,88 | 1 | 1 | 1 | 1 | 1 | 1 | 1 | 1 | 1 | 1 | 1 | 1 | 1 | 1 | 1 | 1 | 1 |
| 11 | 22,34 | 9 | 23,13 | 1 | 1 | 1 | 1 | 1 | 1 | 1 | 1 | 1 | 1 | 1 | 1 | 1 | 1 | 1 | 1 | 1 |
| 16 | 22,55 | 11 | 23,13 | 1 | 1 | 1 | 1 | 1 | 1 | 1 | 1 | 1 | 1 | 1 | 1 | 1 | 1 | 1 | 1 | 1 |
| 10 | 22,88 | 10 | 23,21 | 1 | 1 | 1 | 1 | 1 | 1 | 1 | 1 | 1 | 1 | 1 | 1 | 1 | 1 | 1 | 1 | 1 |
| 13 | 23,32 | 15 | 24,38 | 1 | 1 | 1 | 1 | 1 | 1 | 0 | 1 | 0 | 1 | 1 | 1 | 1 | 1 | 1 | 1 | 1 |
| 17 | 24,00 | 23 | 24,45 | 1 | 1 | 1 | 1 | 1 | 1 | 1 | 1 | 0 | 1 | 1 | 0 | 1 | 0 | 0 | 1 | 1 |
| 23 | 24,04 | 17 | 24,81 | 1 | 1 | 1 | 1 | 1 | 1 | 1 | 1 | 1 | 1 | 1 | 1 | 1 | 1 | 1 | 1 | 0 |
| 15 | 24,14 | 14 | 24,97 | 1 | 1 | 1 | 1 | 1 | 1 | 1 | 1 | 1 | 1 | 1 | 1 | 1 | 1 | 1 | 1 | 0 |
| 14 | 24,28 | 25 | 25,07 | 1 | 1 | 1 | 1 | 1 | 1 | 0 | 1 | 1 | 1 | 1 | 1 | 1 | 1 | 1 | 1 | 0 |
| 27 | 25,14 | 24 | 25,20 | 0 | 1 | 1 | 1 | 1 | 1 | 0 | 1 | 0 | 1 | 1 | 0 | 1 | 1 | 0 | 1 | 0 |
| 24 | 25,24 | 13 | 25,29 | 1 | 1 | 1 | 1 | 1 | 1 | 1 | 1 | 1 | 1 | 1 | 1 | 0 | 1 | 1 | 1 | 0 |
| 31 | 25,27 | 19 | 25,45 | 1 | 1 | 1 | 1 | 1 | 1 | 0 | 1 | 1 | 1 | 1 | 1 | 0 | 1 | 0 | 1 | 0 |
| 18 | 25,30 | 21 | 25,95 | 0 | 1 | 0 | 1 | 1 | 1 | 0 | 1 | 0 | 1 | 0 | 0 | 1 | 1 | 0 | 1 | 1 |
| 26 | 25,47 | 27 | 26,12 | 0 | 1 | 0 | 1 | 1 | 1 | 0 | 1 | 1 | 1 | 1 | 1 | 1 | 1 | 0 | 1 | 1 |
| 19 | 25,50 | 31 | 26,24 | 0 | 1 | 0 | 1 | 1 | 1 | 0 | 0 | 0 | 0 | 0 | 0 | 1 | 0 | 0 | 1 | 0 |
| 21 | 25,54 | 26 | 26,32 | 0 | 1 | 1 | 1 | 1 | 1 | 0 | 1 | 0 | 0 | 0 | 1 | 0 | 0 | 0 | 1 | 0 |
| 22 | 25,87 | 32 | 26,64 | 0 | 1 | 0 | 1 | 1 | 1 | 0 | 1 | 0 | 0 | 0 | 1 | 0 | 1 | 0 | 1 | 0 |
| 20 | 26,27 | 35 | 26,66 | 0 | 1 | 0 | 1 | 1 | 1 | 0 | 0 | 0 | 0 | 0 | 0 | 0 | 1 | 0 | 1 | 0 |
| 32 | 26,44 | 36 | 27,05 | 0 | 1 | 0 | 0 | 1 | 1 | 1 | 1 | 0 | 0 | 0 | 0 | 0 | 0 | 0 | 1 | 0 |
| 35 | 27,04 | 30 | 27,24 | 0 | 1 | 0 | 1 | 1 | 1 | 0 | 1 | 0 | 0 | 0 | 0 | 1 | 0 | 0 | 1 | 0 |
| 28 | 27,14 | 29 | 27,34 | 1 | 1 | 0 | 0 | 1 | 0 | 0 | 1 | 0 | 0 | 0 | 0 | 0 | 1 | 0 | 1 | 0 |
| 29 | 27,15 | 28 | 27,41 | 0 | 1 | 0 | 1 | 1 | 1 | 1 | 0 | 0 | 0 | 0 | 0 | 0 | 0 | 0 | 1 | 0 |
| 40 | 27,65 | 22 | 27,42 | 1 | 1 | 0 | 0 | 1 | 0 | 0 | 1 | 1 | 1 | 1 | 0 | 0 | 1 | 0 | 1 | 0 |
| 34 | 27,89 | 34 | 27,82 | 0 | 1 | 0 | 1 | 0 | 1 | 0 | 0 | 0 | 0 | 0 | 0 | 0 | 1 | 0 | 1 | 0 |
| 36 | 28,13 | 40 | 28,19 | 0 | 0 | 0 | 1 | 0 | 1 | 0 | 0 | 0 | 0 | 0 | 0 | 0 | N/A | 0 | 0 | 0 |
| 38 | 28,14 | 18 | 28,33 | 1 | 1 | 1 | 1 | 1 | 1 | 0 | 1 | 1 | 1 | 1 | 1 | 0 | 1 | 1 | 1 | 0 |
| 42 | 28,43 | 33 | 28,92 | 0 | 1 | 0 | 0 | 0 | 0 | 0 | 0 | 0 | 0 | 0 | 0 | 0 | N/A | 0 | 0 | 0 |
| 30 | 28,86 | 38 | 29,36 | 0 | 1 | 0 | 1 | 1 | 1 | 0 | 0 | 0 | 0 | 0 | 0 | 0 | 0 | 0 | 1 | 0 |
| 33 | 28,96 | 20 | 29,46 | 0 | 1 | 1 | 0 | 1 | 0 | 0 | 1 | 0 | 1 | 1 | 1 | 0 | 1 | 0 | 1 | 0 |
| 44 | 29,24 | 42 | 29,48 | 0 | 0 | 0 | 0 | 0 | 0 | 0 | 0 | 0 | 0 | 0 | 0 | 0 | 0 | 0 | 0 | 0 |
| 25 | 29,70 | 44 | 29,51 | 0 | 0 | 0 | 0 | 0 | 1 | 0 | 0 | 0 | 0 | 0 | 0 | 0 | 0 | 0 | 0 | 0 |
| 39 | 29,76 | 39 | 30,12 | 0 | 0 | 0 | 0 | 0 | 1 | 0 | 0 | 0 | 0 | 0 | 0 | 0 | 0 | 0 | 0 | 0 |
| 45 | 30,10 | 37 | 30,13 | 0 | 1 | 0 | 0 | 0 | 0 | 0 | 0 | 0 | 0 | 0 | 0 | 0 | 0 | 0 | 0 | 0 |
| 41 | 30,13 | 41 | 30,14 | 0 | 0 | 0 | 0 | 0 | 1 | 0 | 0 | 0 | 0 | 0 | 0 | 0 | 0 | 0 | 0 | 0 |
| 37 | 30,54 | 45 | 31,19 | 0 | 0 | 0 | 0 | 0 | 0 | 0 | 0 | 0 | 0 | 0 | 0 | 0 | 0 | 0 | 0 | 0 |
| 43 | 31,05 | 48 | 31,19 | 0 | 1 | 0 | 0 | 0 | 0 | 0 | 0 | 0 | 0 | 0 | 0 | 0 | 0 | 0 | 0 | 0 |
| 46 | 31,54 | 46 | 31,34 | 0 | 1 | 0 | 0 | 0 | 0 | 1 | 0 | 0 | 1 | 0 | 0 | 0 | 0 | 0 | 0 | 0 |
| 48 | 32,06 | 43 | 31,61 | invalid | 0 | 0 | 0 | 0 | 0 | 0 | 0 | 0 | 0 | 0 | 0 | 0 | 0 | 0 | 0 | 0 |
| 47 | 35,19 | 47 | 34,55 | 0 | 0 | 0 | 0 | 0 | 0 | 0 | 0 | 0 | 0 | 0 | 0 | 0 | 0 | 0 | 0 | 0 |
| 49 | 35,22 | 50 | 35,83 | 0 | 0 | 0 | 0 | 0 | 0 | 0 | 0 | 0 | 0 | 0 | 0 | 0 | 0 | 0 | 0 | 0 |
| 50 | 36,36 | 49 | 36,04 | 0 | 0 | 0 | 0 | 0 | 0 | 0 | 0 | 0 | 0 | 0 | 0 | 0 | 0 | 0 | 0 | 0 |

Supplemental Table 1

Comparative evaluation results of SARS-CoV-2 antigen RDT passing the sensitivity criteria

(in alphabetical order of manufacturers)

| | | | | 35 | 36 | 37 | 38 | 39 | 40 | 41 | 42 | 43 | 44 | 45 | 46 | 47 | 48 | 49 | 50 | 51 |
|---|---|---|---|---|---|---|---|---|---|---|---|---|---|---|---|---|---|---|---|---|
| | | **Manufacturer** | | Guangdong Hecin Scientific,Inc. | Guangdong Wesail Biotech Co., Ltd. | Guangzhou Wondfo Biotech Co. Ltd | Hangzhou Clongene Biotech Co., Ltd. | Hangzhou Immuno Biotech Co.,Ltd. | Hangzhou Laihe Biotech Co., Ltd. (Lissner Qi GmbH) | Hangzhou Lysun Biotechnology Co., Ltd. | Hangzhou Testsea Biotechnology Co., Ltd | Humasis Co., Ltd. | IVC Pragen Healthcare | Jiangsu Diagnostics Biotechnology Co., Ltd | Jiangsu Medomics Medical Technology Co., Ltd | Joinstar Biomedical Technology Co., Ltd (CIV care impuls Vertrieb) | Labnovation Technologies, Inc. | Lumigenex (Suzhou) Co., Ltd. | LumiQuick Diagnostics Inc. | LumiraDX |
| | | **Test name** | | 2019-nCoV Antigen Test Kit (colloidal gold method) | COVID-19 Ag Test Kit | Wondfo SARS-CoV-2 Antigen Test (Lateral Flow Method) | Clungene COVID-19 Antigen Rapid Test | IMMUNOBIO SARS-CoV-2 Antigen-Schnelltest (COVID-19 Ag) | Lyher Novel Coronavirus (COVID-19) Antigen Test Kit (Colloidal Gold) | Lysun COVID-19 Antigen Rapid Test Device (Colloidal Gold) | Testsealabs® Rapid Test Kit COVID-19 Antigen Test Cassette | COVID-19 Ag Test | GenBody COVID-19 Ag | COVID-19 Antigen Rapid Test Cassette (Colloidal Gold) | SARS-CoV-2-Antigen-Testkit (LFIA) | COVID-19 Antigen Schnelltest (Colloidal Gold) | Labnovation SARS-CoV-2 Antigen Rapid Test Kit (Immunochromatography) | PocRoc SARS-CoV-2, Antigen Schnelltest Set (Kolloidales Gold) | QuickProfile Covid-19 Antigen Test Card | LumiraDx SARS-CoV-2 Ag Test |
| **Panel 1 V1** | | **Panel 1 V2** | | Panel 1 V2 | Panel 1 V1 | Panel 1 V2 | Panel 1 V1 | Panel 1 V2 | Panel 1 V1 | Panel 1 V2 | Panel 1 V2 | Panel 1 V2 | Panel 1 V1 | Panel 1 V2 | Panel 1 V2 | Panel 1 V1 | Panel 1 V2 | Panel 1 V2 | Panel 1 V2 | Panel 1 V1 |
| **Pool Nr.** | **CT** | **Pool Nr.** | **CT** | | | | | | | | | | | | | | | | | |
| 1 | 17,55 | 1 | 17,31 | 1 | 1 | 1 | 1 | 1 | 1 | 1 | 1 | 1 | 1 | 1 | 1 | 1 | 1 | 1 | 1 | 1 |
| 6 | 20,20 | 2 | 19,08 | 1 | 1 | 1 | 1 | 1 | 1 | 1 | 1 | 1 | 1 | 1 | 1 | 1 | 1 | 1 | 1 | 1 |
| 5 | 20,28 | 3 | 19,62 | 1 | 1 | 1 | 1 | 1 | 1 | 1 | 1 | 1 | 1 | 1 | 1 | 1 | 1 | 1 | 1 | 1 |
| 3 | 20,38 | 5 | 20,60 | 1 | 1 | 1 | 1 | 1 | 1 | 1 | 1 | 1 | 1 | 1 | 1 | 1 | 1 | 1 | 1 | 1 |
| 2 | 20,54 | 4 | 20,61 | 1 | 1 | 1 | 1 | 1 | 1 | 1 | 1 | 1 | 1 | 1 | 1 | 1 | 1 | 1 | 1 | 1 |
| 4 | 20,98 | 6 | 21,21 | 1 | 1 | 1 | 1 | 1 | 1 | 1 | 1 | 1 | 1 | 1 | 1 | 1 | 1 | 1 | 1 | 1 |
| 7 | 21,71 | 12 | 22,12 | 1 | 1 | 1 | 1 | 1 | 1 | 1 | 1 | 1 | 1 | 1 | 1 | 1 | 1 | 1 | 1 | 1 |
| 12 | 21,82 | 7 | 22,15 | 1 | 1 | 1 | 1 | 1 | 1 | 1 | 1 | 1 | 1 | 1 | 1 | 1 | 1 | 1 | 1 | 1 |
| 8 | 21,95 | 8 | 22,32 | 1 | 1 | 1 | 1 | 1 | 1 | 1 | 1 | 1 | 1 | 1 | 1 | 1 | 1 | 1 | 1 | 1 |
| 9 | 22,14 | 16 | 22,88 | 1 | 1 | 1 | 1 | 1 | 1 | 1 | 1 | 1 | 1 | 1 | 1 | 1 | 1 | 1 | 1 | 1 |
| 11 | 22,34 | 9 | 23,13 | 1 | 1 | 1 | 1 | 1 | 1 | 1 | 1 | 1 | 1 | 1 | 1 | 1 | 1 | 1 | 1 | 1 |
| 16 | 22,55 | 11 | 23,13 | 0 | 1 | 1 | 1 | 0 | 1 | 1 | 1 | 0 | 1 | 1 | 1 | 1 | 1 | 1 | 1 | 1 |
| 10 | 22,88 | 10 | 23,21 | 1 | 1 | 1 | 1 | 1 | 1 | 1 | 1 | 1 | 1 | 1 | 1 | 1 | 1 | 1 | 1 | 1 |
| 13 | 23,32 | 15 | 24,38 | 0 | 1 | 1 | 1 | 1 | 1 | 1 | 1 | 1 | 1 | 1 | 1 | 1 | 1 | 1 | 1 | 1 |
| 17 | 24,00 | 23 | 24,45 | 0 | 1 | 0 | 1 | 0 | 1 | 1 | 1 | 0 | 1 | 1 | 0 | 1 | 0 | 1 | 1 | 1 |
| 23 | 24,04 | 17 | 24,81 | 1 | 1 | 1 | 0 | 1 | 0 | 1 | 1 | 1 | 0 | 1 | 1 | 1 | 1 | 1 | 1 | 1 |
| 15 | 24,14 | 14 | 24,97 | 1 | 1 | 0 | 1 | 1 | 1 | 1 | 1 | 1 | 1 | 1 | 1 | 1 | 1 | 1 | 1 | 1 |
| 14 | 24,28 | 25 | 25,07 | 0 | 1 | 0 | 1 | 0 | 1 | 1 | 1 | 1 | 1 | 1 | 0 | 1 | 0 | 1 | 1 | 1 |
| 27 | 25,14 | 24 | 25,20 | 0 | 1 | 0 | 1 | 0 | 0 | 1 | 1 | 0 | 1 | 1 | 0 | 1 | 0 | 1 | 1 | 1 |
| 24 | 25,24 | 13 | 25,29 | 1 | 1 | 0 | 0 | 1 | 0 | 1 | 1 | 1 | 0 | 1 | 1 | 1 | 1 | 1 | 1 | 0 |
| 31 | 25,27 | 19 | 25,45 | 0 | 0 | 0 | 0 | 1 | 0 | 1 | 1 | 0 | 0 | 1 | 1 | 1 | 1 | 1 | 1 | 0 |
| 18 | 25,30 | 21 | 25,95 | 0 | 1 | 0 | 1 | 0 | 1 | 1 | 0 | 1 | 1 | 1 | 0 | 1 | 0 | 0 | 1 | 1 |
| 26 | 25,47 | 27 | 26,12 | 0 | 1 | 0 | 1 | 0 | 1 | 1 | 1 | 1 | 1 | 1 | 0 | 1 | 0 | 1 | 1 | 1 |
| 19 | 25,50 | 31 | 26,24 | 0 | 1 | 0 | 1 | 0 | 1 | 1 | 0 | 0 | 1 | 1 | 0 | 1 | 0 | 1 | 1 | 1 |
| 21 | 25,54 | 26 | 26,32 | 0 | 1 | 0 | 0 | 0 | 0 | 1 | 0 | 0 | 0 | 1 | 0 | 1 | 0 | 1 | 1 | 0 |
| 22 | 25,87 | 32 | 26,64 | 0 | 1 | 0 | 1 | 0 | 1 | 1 | 0 | 0 | 1 | 1 | 0 | 1 | 0 | 1 | 1 | 1 |
| 20 | 26,27 | 35 | 26,66 | 0 | 1 | 0 | 1 | 0 | 0 | 1 | 1 | 0 | 0 | 1 | 0 | 1 | 0 | 1 | 1 | 1 |
| 32 | 26,44 | 36 | 27,05 | 0 | 0 | 0 | 0 | 0 | 0 | 1 | 0 | 0 | 0 | 1 | 0 | 1 | 0 | 1 | 1 | 1 |
| 35 | 27,04 | 30 | 27,24 | 0 | 1 | 0 | 0 | 0 | 0 | 1 | 1 | 0 | 1 | 1 | 0 | 1 | 0 | 0 | 1 | 1 |
| 28 | 27,14 | 29 | 27,34 | 0 | 0 | 0 | 0 | 0 | 0 | 1 | 1 | 0 | 0 | 1 | 0 | 0 | 0 | 1 | 1 | 0 |
| 29 | 27,15 | 28 | 27,41 | 0 | 1 | 0 | 1 | 0 | 0 | 1 | 0 | 0 | 0 | 1 | 0 | 0 | 0 | 0 | 1 | 1 |
| 40 | 27,65 | 22 | 27,42 | 0 | 0 | 0 | 0 | 0 | 0 | 1 | 1 | 0 | 0 | 1 | 1 | 0 | 1 | 1 | 1 | 0 |
| 34 | 27,89 | 34 | 27,82 | 0 | 0 | 0 | 0 | 0 | 0 | 0 | 0 | 0 | 0 | 0 | 0 | 0 | 0 | 0 | 1 | 0 |
| 36 | 28,13 | 40 | 28,19 | 0 | 1 | 0 | 0 | 0 | 0 | 0 | 0 | 0 | 0 | 0 | 0 | 1 | N/A | 0 | 0 | 1 |
| 38 | 28,14 | 18 | 28,33 | 1 | 0 | 0 | 0 | 1 | 0 | 1 | 1 | 1 | 0 | 1 | 1 | 0 | 1 | 1 | 1 | 0 |
| 42 | 28,43 | 33 | 28,92 | 0 | 0 | 0 | 0 | 0 | 0 | 0 | 0 | 0 | 0 | 0 | 0 | 0 | N/A | 0 | 0 | 0 |
| 30 | 28,86 | 38 | 29,36 | 0 | 1 | 0 | 0 | 0 | 0 | 1 | 0 | 0 | 0 | 1 | 0 | 1 | 0 | 1 | 1 | 1 |
| 33 | 28,96 | 20 | 29,46 | 1 | 0 | 0 | 0 | 0 | 0 | 1 | 1 | 0 | 0 | 1 | 1 | 0 | 0 | 1 | 1 | 0 |
| 44 | 29,24 | 42 | 29,48 | 0 | 0 | 0 | 0 | 0 | 0 | 0 | 0 | 0 | 0 | 0 | 0 | 0 | 0 | 0 | 1 | 0 |
| 25 | 29,70 | 44 | 29,51 | 0 | 0 | 0 | 1 | 0 | 0 | 0 | 0 | 0 | 0 | 0 | 0 | 1 | 0 | 0 | 1 | 1 |
| 39 | 29,76 | 39 | 30,12 | 0 | 0 | 0 | 0 | 0 | 0 | 0 | 0 | 0 | 0 | 0 | 0 | 0 | 0 | 0 | 1 | 0 |
| 45 | 30,10 | 37 | 30,13 | 0 | 0 | 0 | 0 | 0 | 0 | 0 | 0 | 0 | 0 | 0 | 0 | 0 | 0 | 0 | 0 | 0 |
| 41 | 30,13 | 41 | 30,14 | 0 | 0 | 0 | 0 | 0 | 0 | 0 | 0 | 0 | 0 | 0 | 0 | 0 | 0 | 0 | 0 | 0 |
| 37 | 30,54 | 45 | 31,19 | 0 | 0 | 0 | 0 | 0 | 0 | 0 | 0 | 0 | 0 | 0 | 0 | 0 | 0 | 0 | 0 | 0 |
| 43 | 31,05 | 48 | 31,19 | 0 | 0 | 0 | 0 | 0 | 0 | 0 | 0 | 0 | 0 | 0 | 0 | 0 | 0 | 0 | 0 | 0 |
| 46 | 31,54 | 46 | 31,34 | 0 | 0 | 0 | 0 | 0 | 0 | 0 | 0 | 0 | 0 | 0 | 0 | 0 | 0 | 0 | 0 | 0 |
| 48 | 32,06 | 43 | 31,61 | 0 | 0 | 0 | 0 | 0 | 0 | 0 | 0 | 0 | 0 | 0 | 0 | 0 | 0 | 0 | 1 | 0 |
| 47 | 35,19 | 47 | 34,55 | 0 | 1 | 0 | 0 | 0 | 0 | 0 | 0 | 0 | 0 | 0 | 0 | 0 | 0 | 0 | 0 | 0 |
| 49 | 35,22 | 50 | 35,83 | 0 | 0 | 0 | 0 | 0 | 0 | 0 | 0 | 0 | 0 | 0 | 0 | 0 | 0 | 0 | 0 | 0 |
| 50 | 36,36 | 49 | 36,04 | 0 | 0 | 0 | 0 | 0 | 0 | 0 | 0 | 0 | 0 | 0 | 0 | 0 | 0 | 0 | 0 | 0 |

Supplemental Table 1

Comparative evaluation results of SARS-CoV-2 antigen RDT passing the sensitivity criteria

(in alphabetical order of manufacturers)

| | | | | 52 | 53 | 54 | 55 | 56 | 57 | 58 | 59 | 60 | 61 | 62 | 63 | 64 | 65 | 66 | 67 | 68 |
|---|---|---|---|---|---|---|---|---|---|---|---|---|---|---|---|---|---|---|---|---|
| | | **Manufacturer** | | MEDsan GmbH | Merlin Biomedical (Xiamen) Co., Ltd. | Mölab GmbH | MP Biomedicals Germany GmbH | nal von minden GmbH | NanoEntek Inc. | Nanjing Norman Biological Technology Co.,Ltd | Nantong Diagnos Biotechnology Co., Ltd. | New Gene (Hangzhou) Bioengineering Co., Ltd. | Novatech Tibbi Cihaz Ürünleri San. Ve Tic. A.S. | Oncosem Onkolojik Sistemler San. Ve Tic. A.S. | PCL, Inc. | PerGrande BioTech Development Co., Ltd. | Precision Biosensor Inc. (Axon Lab AG) | ProGnosis Biotech | Quidel Corporation | Qingdao Hightop Biotech Co., Ltd. |
| | | **Test name** | | MEDsan® SARS-CoV-2 Antigen Rapid Test | SARS-CoV-2 Antigen Rapid Test Cassette | mö-screen Corona Antigen Test | Rapid SARS-CoV-2 Antigen Test Card | NADAL® COVID-19 Ag Schnelltest | Frend™ COVID-19 Ag | Novel Coronavirus (2019-nCOV) Antigen Testing Kit (Colloidal Gold) | COVID-19 Antigen Saliva Test Kit (Colloidal Gold) | Covid-19-Antigen-Testkit | novacheck®-Ag SARS-CoV-2 Covid-19 Antigen Rapid Test | CAT Antigen Covid Rapid Test | PCL COVID19 Ag Gold Saliva | SARS-CoV-2 Antigen Detection Kit (Colloidal Gold Immuno-chromatographic Assay) | Exdia COVID-19-Ag-Test | Rapid Test Ag 2019-nCoV | Sofia SARS Antigen FIA | Hightop SARS-CoV-2 (Covid-19) Antigen Rapid Test |
| **Panel 1 V1** | | **Panel 1 V2** | | Panel 1 V1 | Panel 1 V2 | Panel 1 V1 | Panel 1 V2 | Panel 1 V1 | Panel 1 V2 | Panel 1 V2 | Panel 1 V2 | Panel 1 V2 | Panel 1 V2 | Panel 1 V2 | Panel 1 V2 | Panel 1 V2 | Panel 1 V1 | Panel 1 V2 | Panel 1 V1 | Panel 1 V2 |
| **Pool Nr.** | **CT** | **Pool Nr.** | **CT** | | | | | | | | | | | | | | | | | |
| 1 | 17,55 | 1 | 17,31 | 1 | 1 | 1 | 1 | 1 | 1 | 1 | 1 | 1 | 1 | 1 | 1 | 1 | 1 | 1 | 1 | 1 |
| 6 | 20,20 | 2 | 19,08 | 1 | 1 | 1 | 1 | 1 | 1 | 1 | 1 | 1 | 1 | 1 | 1 | 1 | 1 | 1 | 1 | 1 |
| 5 | 20,28 | 3 | 19,62 | 1 | 1 | 1 | 1 | 1 | 1 | 1 | 1 | 1 | 1 | 1 | 1 | 1 | 1 | 1 | 1 | 1 |
| 3 | 20,38 | 5 | 20,60 | 1 | 1 | 1 | 1 | 1 | 1 | 1 | 1 | 1 | 1 | 1 | 1 | 1 | 1 | 1 | 1 | 1 |
| 2 | 20,54 | 4 | 20,61 | 1 | 1 | 1 | 1 | 1 | 1 | 1 | 1 | 1 | 1 | 1 | 1 | 1 | 1 | 1 | 1 | 1 |
| 4 | 20,98 | 6 | 21,21 | 1 | 1 | 1 | 1 | 1 | 1 | 1 | 1 | 1 | 1 | 1 | 1 | 1 | 1 | 1 | 1 | 1 |
| 7 | 21,71 | 12 | 22,12 | 1 | 1 | 1 | 1 | 1 | 1 | 1 | 1 | 1 | 1 | 1 | 1 | 1 | 1 | 1 | 1 | 1 |
| 12 | 21,82 | 7 | 22,15 | 1 | 1 | 1 | 1 | 1 | 1 | 1 | 1 | 1 | 1 | 1 | 1 | 1 | 1 | 1 | 1 | 1 |
| 8 | 21,95 | 8 | 22,32 | 1 | 1 | 1 | 1 | 1 | 1 | 1 | 1 | 1 | 1 | 1 | 1 | 1 | 1 | 1 | 1 | 1 |
| 9 | 22,14 | 16 | 22,88 | 1 | 1 | 1 | 1 | 1 | 1 | 1 | 1 | 1 | 1 | 1 | 1 | 1 | 1 | 0 | 1 | 1 |
| 11 | 22,34 | 9 | 23,13 | 1 | 1 | 1 | 1 | 1 | 1 | 1 | 1 | 1 | 1 | 1 | 1 | 1 | 1 | 1 | 1 | 1 |
| 16 | 22,55 | 11 | 23,13 | 1 | 1 | 1 | 1 | 1 | 1 | 1 | 1 | 1 | 1 | 1 | 1 | 1 | 1 | 1 | 1 | 1 |
| 10 | 22,88 | 10 | 23,21 | 1 | 1 | 1 | 1 | 1 | 1 | 1 | 1 | 1 | 1 | 1 | 1 | 1 | 1 | 1 | 1 | 1 |
| 13 | 23,32 | 15 | 24,38 | 1 | 1 | 1 | 1 | 0 | 0 | 1 | 1 | 1 | 1 | 0 | 1 | 1 | 1 | 1 | 1 | 1 |
| 17 | 24,00 | 23 | 24,45 | 1 | 1 | 1 | 1 | 1 | 0 | 0 | 1 | 1 | 0 | 1 | 1 | 1 | 1 | 1 | 1 | 1 |
| 23 | 24,04 | 17 | 24,81 | 1 | 1 | 1 | 1 | 0 | 1 | 1 | 1 | 1 | 1 | 1 | 1 | 1 | 1 | 1 | 0 | 1 |
| 15 | 24,14 | 14 | 24,97 | 1 | 1 | 1 | 1 | 1 | 1 | 1 | 1 | 1 | 1 | 1 | 1 | 1 | 1 | 1 | 0 | 1 |
| 14 | 24,28 | 25 | 25,07 | 1 | 1 | 1 | 1 | 0 | 0 | 1 | 1 | 1 | 1 | 1 | 1 | 0 | 1 | 1 | 1 | 1 |
| 27 | 25,14 | 24 | 25,20 | 1 | 1 | 1 | 1 | 0 | 0 | 0 | 1 | 1 | 0 | 1 | 1 | 0 | 1 | 1 | 0 | 1 |
| 24 | 25,24 | 13 | 25,29 | 0 | 1 | 1 | 1 | 0 | 1 | 1 | 1 | 1 | 1 | 1 | 1 | 0 | 1 | 1 | 0 | 1 |
| 31 | 25,27 | 19 | 25,45 | 0 | 1 | 0 | 1 | 0 | 0 | 1 | 1 | 1 | 0 | 1 | 1 | 0 | 1 | 1 | 0 | 1 |
| 18 | 25,30 | 21 | 25,95 | 1 | 1 | 1 | 1 | 0 | 0 | 0 | 1 | 1 | 0 | 0 | 1 | 0 | 1 | 1 | 1 | 0 |
| 26 | 25,47 | 27 | 26,12 | 1 | 1 | 1 | 1 | 1 | 0 | 1 | 1 | 1 | 0 | 1 | 1 | 0 | 1 | 0 | 1 | 1 |
| 19 | 25,50 | 31 | 26,24 | 1 | 1 | 1 | 0 | 0 | 0 | 0 | 0 | 1 | 0 | 0 | 0 | 0 | 1 | 1 | 0 | 0 |
| 21 | 25,54 | 26 | 26,32 | 1 | 1 | 1 | 1 | 1 | 0 | 0 | 1 | 1 | 0 | 0 | 1 | 1 | 0 | 1 | 0 | 0 |
| 22 | 25,87 | 32 | 26,64 | 1 | 1 | 1 | 0 | 1 | 0 | 0 | 0 | 1 | 0 | 0 | 0 | 0 | 1 | 1 | 0 | 0 |
| 20 | 26,27 | 35 | 26,66 | 1 | 1 | 1 | 0 | 0 | 0 | 0 | 1 | 1 | 0 | 0 | 0 | 0 | 1 | 0 | 0 | 1 |
| 32 | 26,44 | 36 | 27,05 | 1 | 1 | 0 | 1 | 0 | 0 | 0 | 0 | 1 | 0 | 0 | 0 | 1 | 0 | 1 | 0 | 0 |
| 35 | 27,04 | 30 | 27,24 | 1 | 1 | 1 | 0 | 0 | 0 | 0 | 1 | 1 | 1 | 0 | 1 | 0 | 1 | 1 | 0 | 0 |
| 28 | 27,14 | 29 | 27,34 | 0 | 1 | 0 | 0 | 0 | 0 | 0 | 0 | 1 | 0 | 0 | 1 | 0 | 0 | 0 | 0 | 1 |
| 29 | 27,15 | 28 | 27,41 | 0 | 1 | 1 | 0 | 0 | 0 | 0 | 0 | 1 | 0 | 0 | 0 | 1 | 1 | 1 | 0 | 0 |
| 40 | 27,65 | 22 | 27,42 | 0 | 1 | 0 | 1 | 0 | 0 | 1 | 1 | 1 | 1 | 1 | 1 | 0 | 0 | 1 | 0 | 1 |
| 34 | 27,89 | 34 | 27,82 | 0 | 1 | 0 | 0 | 0 | 0 | 0 | 0 | 1 | 0 | 0 | 0 | 0 | 0 | 0 | 0 | 0 |
| 36 | 28,13 | 40 | 28,19 | 0 | 0 | 0 | 0 | 0 | 0 | 0 | 0 | 0 | 0 | 0 | 0 | 0 | 1 | 0 | 0 | 0 |
| 38 | 28,14 | 18 | 28,33 | 1 | 1 | 0 | 1 | 0 | 1 | 1 | 1 | 1 | 1 | 1 | 1 | 0 | 1 | 1 | 0 | 1 |
| 42 | 28,43 | 33 | 28,92 | 1 | 0 | 0 | 0 | 0 | 0 | 0 | 0 | 0 | 0 | 0 | 0 | 1 | 0 | 1 | 0 | 0 |
| 30 | 28,86 | 38 | 29,36 | 0 | 1 | 0 | 0 | 0 | 0 | 0 | 1 | 1 | 0 | 0 | 0 | 0 | 1 | 0 | 0 | 0 |
| 33 | 28,96 | 20 | 29,46 | 0 | 1 | 0 | 0 | 0 | 0 | 0 | 1 | 1 | 0 | 0 | 1 | 0 | 0 | 1 | 0 | 1 |
| 44 | 29,24 | 42 | 29,48 | 0 | 0 | 0 | 0 | 0 | 0 | 0 | 0 | 1 | 0 | 0 | 0 | 0 | 0 | 0 | 0 | 0 |
| 25 | 29,70 | 44 | 29,51 | 0 | 0 | 1 | 0 | 0 | 0 | 0 | 0 | 0 | 0 | 0 | 0 | 0 | 1 | 0 | 0 | 0 |
| 39 | 29,76 | 39 | 30,12 | 0 | 0 | 0 | 0 | 0 | 0 | 0 | 0 | 0 | 0 | 0 | 0 | 0 | 0 | 0 | 0 | 0 |
| 45 | 30,10 | 37 | 30,13 | 0 | 0 | 0 | 0 | 0 | 0 | 0 | 0 | 0 | 0 | 0 | 0 | 0 | 0 | 1 | 0 | 0 |
| 41 | 30,13 | 41 | 30,14 | 0 | 0 | 0 | 0 | 0 | 0 | 0 | 0 | 1 | 0 | 0 | 0 | 0 | 0 | 0 | 0 | 0 |
| 37 | 30,54 | 45 | 31,19 | 0 | 0 | 0 | 0 | 0 | 0 | 0 | 0 | 0 | 0 | 0 | 0 | 0 | 0 | 0 | 0 | 0 |
| 43 | 31,05 | 48 | 31,19 | 0 | 0 | 0 | 0 | 0 | 0 | 0 | 0 | 0 | 0 | 0 | 0 | 0 | 0 | 0 | 0 | 0 |
| 46 | 31,54 | 46 | 31,34 | 0 | 0 | 0 | 0 | 0 | 0 | 0 | 0 | 0 | 0 | 0 | 0 | 0 | 0 | 0 | 0 | 0 |
| 48 | 32,06 | 43 | 31,61 | 0 | 0 | 0 | 0 | 0 | 0 | 0 | 0 | 1 | 0 | 0 | 0 | 0 | 0 | 0 | 0 | 0 |
| 47 | 35,19 | 47 | 34,55 | 0 | 0 | 0 | 0 | 0 | 0 | 0 | 0 | 0 | 0 | 0 | 0 | 0 | 0 | 0 | 0 | 0 |
| 49 | 35,22 | 50 | 35,83 | 0 | 0 | 0 | 0 | 0 | 0 | 0 | 0 | 0 | 0 | 0 | 0 | 0 | 0 | 0 | 0 | 0 |
| 50 | 36,36 | 49 | 36,04 | 0 | 0 | 0 | 0 | 0 | 0 | 0 | 0 | 0 | 0 | 0 | 0 | 0 | 0 | 0 | 0 | 0 |

Supplemental Table 1

Comparative evaluation results of SARS-CoV-2 antigen RDT passing the sensitivity criteria

(in alphabetical order of manufacturers)

| | | | | 69 | 70 | 71 | 72 | 73 | 74 | 75 | 76 | 77 | 78 | 79 | 80 | 81 | 82 | 83 | 84 | 85 |
|---|---|---|---|---|---|---|---|---|---|---|---|---|---|---|---|---|---|---|---|---|
| | | **Manufacturer** | | R-Biopharm AG | Safecare Biotech Hangzhou Co., Ltd. | Salofa OY | ScheBo Biotech AG | SD BIOSENSOR (Roche Diagnostics GmbH) | SD BIOSENSOR | SD BIOSENSOR | SGA Mühendislik DAN. EG. Icve DIS.Ltd.STI | Shenzhen Lvshiyuan Biotechnology Co., Ltd. | Shenzhen Microprofit Biotech Co., Ltd. | Shenzhen Watmind Medical Co.,Ltd. | Shenzhen Watmind Medical Co.,Ltd. | Shenzhen Zhenrui Biotech co.Ltd. | Siemens Healthineers | Sugentech, Inc. | Toda Pharma | Triplex International Biosciences (China) Co., Ltd. |
| | | **Test name** | | RIDA® QUICK SARS-CoV-2 Antigen | Safecare COVID-19 Ag Rapid Test Kit (Swab) | salocor SARS-CoV-2 Antigen Rapid Test Cassette (Nasopharyngeal swab) | ScheBo SARS-CoV-2 Quick Antigen | SARS-CoV-2 Rapid Antigen Test | STANDARD™ F COVID-19 Ag FIA | STANDARD™ Q COVID-19 Ag Test | V-Chek SARS-CoV-2 Rapid Ag Test Kit (Collaidal Gold) | Green Spring SARS-CoV-2 Antigen Rapid Test Kit (Colloidal Gold) | fluorecare COVID-19 SARS-CoV-2 Spike Protein Test Kit (Colloidal Gold Chromatographic Immunoassay) | SARS-CoV-2 Ag Diagnostic Test Kit (Colloidal Gold) | SARS-CoV-2 Ag Diagnostic Test Kit (Immuno-fluorescence) | Zhenrui COVID-19 (SARS-COV-2) Antigen Test Kits | CLINITEST® Rapid COVID-19 Antigen Test | SGTi-flex COVID-19 Ag | Toda Coronadiag Ag | SARS-CoV-2 Antigen Rapid Test Kit |
| **Panel 1 V1** | | **Panel 1 V2** | | Panel 1 V1 | Panel 1 V2 | Panel 1 V2 | Panel 1 V2 | Panel 1 V1 | Panel 1 V1 | Panel 1 V1 | Panel 1 V2 | Panel 1 V2 | Panel 1 V2 | Panel 1 V2 | Panel 1 V2 | Panel 1 V2 | Panel 1 V1 | Panel 1 V2 | Panel 1 V2 | Panel 1 V2 |
| **Pool Nr.** | **CT** | **Pool Nr.** | **CT** | | | | | | | | | | | | | | | | | |
| 1 | 17,55 | 1 | 17,31 | 1 | 1 | 1 | 1 | 1 | 1 | 1 | 1 | 1 | 1 | 1 | 1 | 1 | 1 | 1 | 1 | 1 |
| 6 | 20,20 | 2 | 19,08 | 1 | 1 | 1 | 1 | 1 | 1 | 1 | 1 | 1 | 1 | 1 | 1 | 1 | 1 | 1 | 1 | 1 |
| 5 | 20,28 | 3 | 19,62 | 1 | 1 | 1 | 1 | 1 | 1 | 1 | 1 | 1 | 1 | 1 | 1 | 1 | 1 | 1 | 1 | 1 |
| 3 | 20,38 | 5 | 20,60 | 1 | 1 | 1 | 1 | 1 | 1 | 1 | 1 | 1 | 1 | 1 | 1 | 1 | 1 | 1 | 1 | 1 |
| 2 | 20,54 | 4 | 20,61 | 1 | 1 | 1 | 1 | 1 | 1 | 1 | 1 | 1 | 1 | 1 | 1 | 1 | 1 | 1 | 1 | 1 |
| 4 | 20,98 | 6 | 21,21 | 1 | 1 | 1 | 1 | 1 | 1 | 1 | 1 | 1 | 1 | 1 | 1 | 1 | 1 | 1 | 1 | 1 |
| 7 | 21,71 | 12 | 22,12 | 1 | 1 | 1 | 1 | 1 | 1 | 1 | 1 | 1 | 1 | 1 | 1 | 1 | 1 | 1 | 1 | 1 |
| 12 | 21,82 | 7 | 22,15 | 1 | 1 | 1 | 1 | 1 | 1 | 1 | 1 | 1 | 1 | 1 | 1 | 1 | 1 | 1 | 1 | 1 |
| 8 | 21,95 | 8 | 22,32 | 1 | 1 | 1 | 1 | 1 | 1 | 1 | 1 | 1 | 1 | 1 | 1 | 1 | 1 | 1 | 1 | 1 |
| 9 | 22,14 | 16 | 22,88 | 1 | 1 | 1 | 1 | 1 | 1 | 1 | 1 | 1 | 1 | 1 | 1 | 1 | 1 | 1 | 1 | 1 |
| 11 | 22,34 | 9 | 23,13 | 1 | 1 | 1 | 1 | 1 | 1 | 1 | 1 | 1 | 1 | 1 | 1 | 1 | 1 | 1 | 1 | 1 |
| 16 | 22,55 | 11 | 23,13 | 1 | 1 | 0 | 1 | 1 | 1 | 1 | 1 | 1 | 1 | 1 | 1 | 1 | 1 | 1 | 1 | 1 |
| 10 | 22,88 | 10 | 23,21 | 1 | 1 | 1 | 1 | 1 | 1 | 1 | 1 | 1 | 1 | 1 | 1 | 1 | 1 | 1 | 1 | 1 |
| 13 | 23,32 | 15 | 24,38 | 1 | 1 | 0 | 1 | 1 | 1 | 1 | 1 | 1 | 1 | 1 | 1 | 0 | 1 | 1 | 1 | 1 |
| 17 | 24,00 | 23 | 24,45 | 1 | 1 | 0 | 1 | 1 | 1 | 1 | 0 | 1 | 1 | 1 | 1 | 0 | 1 | 1 | 1 | 1 |
| 23 | 24,04 | 17 | 24,81 | 1 | 1 | 1 | 1 | 0 | 1 | 0 | 1 | 1 | 1 | 1 | 1 | 1 | 1 | 1 | 1 | 1 |
| 15 | 24,14 | 14 | 24,97 | 1 | 1 | 1 | 1 | 0 | 1 | 0 | 1 | 1 | 1 | 1 | 1 | 0 | 1 | 1 | 1 | 1 |
| 14 | 24,28 | 25 | 25,07 | 1 | 1 | 1 | 1 | 1 | 1 | 1 | 1 | 1 | 1 | 1 | 1 | 1 | 1 | 1 | 1 | 1 |
| 27 | 25,14 | 24 | 25,20 | 0 | 1 | 0 | 1 | 1 | 1 | 1 | 0 | 1 | 1 | 1 | 1 | 0 | 1 | 1 | 1 | 1 |
| 24 | 25,24 | 13 | 25,29 | 1 | 1 | 1 | 1 | 0 | 1 | 0 | 1 | 1 | 1 | 1 | 1 | 1 | 1 | 1 | 1 | 1 |
| 31 | 25,27 | 19 | 25,45 | 0 | 1 | 0 | 1 | 0 | 0 | 0 | 1 | 1 | 1 | 1 | 1 | 0 | 1 | 1 | 1 | 1 |
| 18 | 25,30 | 21 | 25,95 | 1 | 1 | 0 | 1 | 1 | 1 | 1 | N/A | 1 | 1 | 1 | 1 | 0 | 1 | 0 | 1 | 1 |
| 26 | 25,47 | 27 | 26,12 | 1 | 1 | 0 | 1 | 1 | 1 | 1 | 0 | 1 | 1 | 1 | 1 | 0 | 1 | 1 | 1 | 1 |
| 19 | 25,50 | 31 | 26,24 | 0 | 0 | 0 | 1 | 1 | 1 | 1 | 0 | 1 | 0 | 1 | 0 | 0 | 1 | 1 | 1 | 1 |
| 21 | 25,54 | 26 | 26,32 | 1 | 1 | 0 | 1 | 0 | 1 | 0 | 0 | 1 | 0 | 1 | 1 | 0 | 1 | 1 | 1 | 1 |
| 22 | 25,87 | 32 | 26,64 | 0 | 0 | 0 | 1 | 1 | 1 | 1 | 0 | 1 | 0 | 1 | 0 | 0 | 1 | 0 | 1 | 1 |
| 20 | 26,27 | 35 | 26,66 | 0 | 1 | 0 | 1 | 0 | 1 | 0 | 0 | 1 | 0 | 1 | 1 | 0 | 1 | 1 | 1 | 1 |
| 32 | 26,44 | 36 | 27,05 | 0 | 1 | 0 | 1 | 0 | 0 | 0 | 0 | 1 | 1 | 1 | 1 | 0 | 1 | 1 | 1 | 1 |
| 35 | 27,04 | 30 | 27,24 | 0 | 1 | 0 | 1 | 1 | 1 | 1 | 0 | 1 | 0 | 1 | 1 | 0 | 1 | 1 | 1 | 1 |
| 28 | 27,14 | 29 | 27,34 | 0 | 1 | 0 | 1 | 0 | 1 | 0 | 0 | 1 | 0 | 1 | 0 | 0 | 1 | 1 | 1 | 1 |
| 29 | 27,15 | 28 | 27,41 | 0 | 0 | 0 | 1 | 0 | 1 | 0 | 0 | 1 | 0 | 1 | 0 | 0 | 1 | 1 | 1 | 1 |
| 40 | 27,65 | 22 | 27,42 | 0 | 1 | 0 | 1 | 0 | 0 | 0 | 1 | 1 | 1 | 1 | 1 | 0 | 0 | 1 | 1 | 1 |
| 34 | 27,89 | 34 | 27,82 | 0 | 0 | 0 | 1 | 0 | 1 | 0 | 0 | 1 | 1 | 1 | 0 | 0 | 1 | 1 | 1 | 1 |
| 36 | 28,13 | 40 | 28,19 | 0 | 0 | N/A | 1 | 1 | 1 | 1 | N/A | 1 | 0 | 1 | 0 | 0 | 1 | 0 | 1 | 0 |
| 38 | 28,14 | 18 | 28,33 | 0 | 1 | 1 | 1 | 0 | 1 | 0 | 1 | 1 | 1 | 1 | 1 | 1 | 1 | 1 | 1 | 1 |
| 42 | 28,43 | 33 | 28,92 | 0 | 0 | 0 | 0 | 0 | 0 | 0 | N/A | 1 | 0 | 1 | 0 | 0 | 1 | 0 | 1 | 0 |
| 30 | 28,86 | 38 | 29,36 | 0 | 1 | 0 | 1 | 0 | 0 | 0 | 0 | 1 | 0 | 1 | 1 | 0 | 1 | 1 | 1 | 1 |
| 33 | 28,96 | 20 | 29,46 | 0 | 0 | 0 | 1 | 0 | 0 | 0 | 1 | 1 | 1 | 1 | 1 | 0 | 1 | 1 | 1 | 1 |
| 44 | 29,24 | 42 | 29,48 | 0 | 0 | 0 | 1 | 0 | 0 | 0 | 0 | 1 | 0 | 1 | 0 | 0 | 0 | 0 | 1 | 1 |
| 25 | 29,70 | 44 | 29,51 | 0 | 0 | 0 | 0 | 0 | 1 | 0 | 0 | 0 | 0 | 0 | 0 | 0 | 1 | 0 | 0 | 0 |
| 39 | 29,76 | 39 | 30,12 | 0 | 0 | 0 | 0 | 0 | 0 | 0 | 0 | 1 | 0 | 0 | 0 | 0 | 0 | 0 | 0 | 0 |
| 45 | 30,10 | 37 | 30,13 | 0 | 0 | 0 | 0 | 0 | 0 | 0 | 0 | 1 | 1 | 1 | 0 | 0 | 0 | 0 | 1 | 0 |
| 41 | 30,13 | 41 | 30,14 | 0 | 0 | 0 | 0 | 0 | 0 | 0 | 0 | 0 | 0 | 1 | 0 | 0 | 0 | 0 | 0 | 1 |
| 37 | 30,54 | 45 | 31,19 | 0 | 0 | 0 | 1 | 0 | 0 | 0 | N/A | 0 | 0 | 0 | 0 | 0 | 0 | 0 | 1 | 0 |
| 43 | 31,05 | 48 | 31,19 | 0 | 0 | 0 | 0 | 0 | 0 | 0 | 0 | 0 | 0 | 0 | 0 | 0 | 0 | 0 | 0 | 0 |
| 46 | 31,54 | 46 | 31,34 | 0 | 0 | 0 | 0 | 0 | 0 | 0 | 0 | 1 | 0 | 0 | 0 | 0 | 0 | 0 | 1 | 0 |
| 48 | 32,06 | 43 | 31,61 | 0 | 0 | 0 | 0 | 0 | 0 | 0 | 0 | 0 | 0 | 0 | 0 | 0 | 0 | 0 | 0 | 1 |
| 47 | 35,19 | 47 | 34,55 | 0 | 0 | 0 | 0 | 0 | 0 | 0 | 0 | 1 | 0 | 0 | 0 | 0 | 0 | 0 | 1 | 0 |
| 49 | 35,22 | 50 | 35,83 | 0 | 0 | 0 | 0 | 0 | 0 | 0 | 0 | 0 | 0 | 0 | 0 | 0 | 0 | 0 | 0 | 0 |
| 50 | 36,36 | 49 | 36,04 | 0 | 0 | 0 | 0 | 0 | 0 | 0 | 0 | 0 | 0 | 0 | 0 | 0 | 0 | 0 | 0 | 0 |

Supplemental Table 1
Comparative evaluation results of SARS-CoV-2 antigen RDT passing the sensitivity criteria
(in alphabetical order of manufacturers)

| | | | | 86 | 87 | 88 | 89 | 90 | 91 | 92 | 93 | 94 | 95 | 96 |
|---|---|---|---|---|---|---|---|---|---|---|---|---|---|---|
| | | **Manufacturer** | | ulti med Products (Deutschland) GmbH | Vitrosens Biyoteknoloji Ltd. Sti | Wantai (Beijing Wantai Biological Pharmacy Enterprise Co., Ltd.) | Wuhan EasyDiagnosis Biomedicine Co., Ltd | Wuhan Life Origin Biotech Joint Stock Co., Ltd. | Wuhan UNscience Biotechnology Co., Ltd. | Xiamen Boson Biotech Co., Ltd. | Xiamen WIZ Biotech Co., Ltd. | Zet Medikal Tekstil Dis Ticaret Ltd. STI. | Zhejiang Anji Saianfu Biotech Co.,Ltd. | Zhejiang Orient Gene Biotech Co.,Ltd |
| | | **Test name** | | COVID-19 Antigen Speicheltest (Immunochromatographie) | RapidFor SARS-CoV-2 Rapid Antigen Test Colloidal Gold | SARS-CoV-2 Ag Rapid Test (FIA) | COVID-19 (SARS-CoV-2) Antigen Test Kit | SARS-CoV-2 Antigen Assay Kit (Immunochromatography) | SARS-CoV-2 Antigen Rapid Test Kit | SARS-CoV-2 Antigen Schnelltest | Wizbiotech SARS-CoV-2 Antigen Rapid Test | softec SARS COV-2 (Covid-19) Antigen Test Kit | reOpenTest COVID-19 Antigen Rapid Test (Colloidal Gold) | Coronavirus Ag Rapid Test Cassette (Swab) |
| **Panel 1 V1** | | **Panel 1 V2** | | Panel 1 V2 | Panel 1 V2 | Panel 1 V1 | Panel 1 V2 | Panel 1 V2 | Panel 1 V2 | Panel 1 V2 | Panel 1 V2 | Panel 1 V2 | Panel 1 V2 | Panel 1 V1 |
| **Pool Nr.** | **CT** | **Pool Nr.** | **CT** | | | | | | | | | | | |
| 1 | 17,55 | 1 | 17,31 | 1 | 1 | 1 | 1 | 1 | 1 | 1 | 1 | 1 | 1 | 1 |
| 6 | 20,20 | 2 | 19,08 | 1 | 1 | 1 | 1 | 1 | 1 | 1 | 1 | 1 | 1 | 1 |
| 5 | 20,28 | 3 | 19,62 | 1 | 1 | 1 | 1 | 1 | 1 | 1 | 1 | 1 | 1 | 1 |
| 3 | 20,38 | 5 | 20,60 | 1 | 1 | 1 | 1 | 1 | 1 | 1 | 1 | 1 | 1 | 1 |
| 2 | 20,54 | 4 | 20,61 | 1 | 1 | 1 | 1 | 1 | 1 | 1 | 1 | 0 | 1 | 1 |
| 4 | 20,98 | 6 | 21,21 | 1 | 1 | 1 | 1 | 1 | 1 | 1 | 1 | 1 | 1 | 1 |
| 7 | 21,71 | 12 | 22,12 | 1 | 1 | 1 | 1 | 1 | 1 | 1 | 1 | 1 | 1 | 1 |
| 12 | 21,82 | 7 | 22,15 | 1 | 1 | 1 | 1 | 1 | 1 | 1 | 1 | 1 | 1 | 1 |
| 8 | 21,95 | 8 | 22,32 | 1 | 1 | 1 | 1 | 1 | 1 | 1 | 1 | 1 | 1 | 1 |
| 9 | 22,14 | 16 | 22,88 | 1 | 1 | 1 | 1 | 1 | 1 | 1 | 1 | 1 | 1 | 1 |
| 11 | 22,34 | 9 | 23,13 | 1 | 1 | 1 | 1 | 1 | 1 | 1 | 1 | 1 | 1 | 1 |
| 16 | 22,55 | 11 | 23,13 | 1 | 1 | 1 | 1 | 1 | 1 | 1 | 1 | 1 | 1 | 1 |
| 10 | 22,88 | 10 | 23,21 | 1 | 1 | 1 | 1 | 1 | 1 | 1 | 1 | 0 | 1 | 1 |
| 13 | 23,32 | 15 | 24,38 | 1 | 1 | 1 | 1 | 1 | 0 | 1 | 0 | 1 | 1 | 1 |
| 17 | 24,00 | 23 | 24,45 | 1 | 1 | 1 | 1 | 1 | 0 | 1 | 0 | N/A | 0 | 1 |
| 23 | 24,04 | 17 | 24,81 | 1 | 1 | 1 | 1 | 1 | 1 | 1 | 1 | 1 | 1 | 1 |
| 15 | 24,14 | 14 | 24,97 | 1 | 1 | 1 | 1 | 1 | 1 | 1 | 1 | 1 | 1 | 1 |
| 14 | 24,28 | 25 | 25,07 | 1 | 1 | 1 | 1 | 1 | 1 | 1 | 0 | 1 | 1 | 1 |
| 27 | 25,14 | 24 | 25,20 | 1 | 0 | 1 | 1 | 1 | 0 | 1 | 0 | 1 | 0 | 1 |
| 24 | 25,24 | 13 | 25,29 | 1 | 1 | 1 | 1 | 1 | 1 | 1 | 1 | 1 | 1 | 1 |
| 31 | 25,27 | 19 | 25,45 | 1 | 1 | 1 | 1 | 1 | 1 | 1 | 1 | 0 | 1 | 1 |
| 18 | 25,30 | 21 | 25,95 | 1 | 0 | 1 | 1 | 1 | 0 | 1 | 0 | N/A | 0 | 1 |
| 26 | 25,47 | 27 | 26,12 | 1 | 1 | 1 | 1 | 1 | 0 | 1 | 0 | 0 | 1 | 1 |
| 19 | 25,50 | 31 | 26,24 | 1 | 0 | 1 | 1 | 1 | 0 | 0 | 0 | invalid | 0 | 1 |
| 21 | 25,54 | 26 | 26,32 | 1 | 0 | 1 | 1 | 0 | 0 | 1 | 0 | 1 | 0 | 1 |
| 22 | 25,87 | 32 | 26,64 | 1 | 0 | 1 | 1 | 1 | 0 | 0 | 0 | 0 | 0 | 1 |
| 20 | 26,27 | 35 | 26,66 | 1 | 0 | 1 | 1 | 1 | 0 | 0 | 0 | 0 | 1 | 1 |
| 32 | 26,44 | 36 | 27,05 | 1 | 0 | 1 | 1 | 0 | 0 | 1 | 0 | 0 | 0 | 1 |
| 35 | 27,04 | 30 | 27,24 | 1 | 0 | 1 | 1 | 1 | 0 | 0 | 0 | 0 | 0 | 1 |
| 28 | 27,14 | 29 | 27,34 | 1 | 0 | 1 | 1 | 0 | 0 | 0 | 0 | 0 | 0 | 1 |
| 29 | 27,15 | 28 | 27,41 | 1 | 0 | 1 | 0 | 0 | 0 | 0 | 0 | 0 | 0 | 1 |
| 40 | 27,65 | 22 | 27,42 | 1 | 1 | 0 | 1 | 1 | 0 | 1 | 0 | 1 | 1 | 0 |
| 34 | 27,89 | 34 | 27,82 | 1 | 0 | 1 | 0 | 0 | 0 | 0 | 0 | 0 | 0 | 1 |
| 36 | 28,13 | 40 | 28,19 | 1 | 0 | 1 | 0 | 0 | 0 | 0 | 0 | N/A | 0 | 1 |
| 38 | 28,14 | 18 | 28,33 | 1 | 1 | 1 | 1 | 1 | 1 | 1 | 1 | N/A | 1 | 1 |
| 42 | 28,43 | 33 | 28,92 | 1 | 0 | 0 | 0 | 0 | 0 | 0 | 0 | N/A | 0 | 1 |
| 30 | 28,86 | 38 | 29,36 | 1 | 0 | 1 | 1 | 0 | 0 | 0 | 0 | 0 | 0 | 1 |
| 33 | 28,96 | 20 | 29,46 | 1 | 1 | 0 | 1 | 1 | 0 | 0 | 0 | 0 | 0 | 1 |
| 44 | 29,24 | 42 | 29,48 | 1 | 0 | 0 | 0 | 0 | 0 | 0 | 0 | 0 | 0 | 0 |
| 25 | 29,70 | 44 | 29,51 | 0 | 0 | 1 | 0 | 0 | 0 | 0 | 0 | 0 | 0 | 1 |
| 39 | 29,76 | 39 | 30,12 | 0 | 0 | 0 | 0 | 0 | 0 | 0 | 0 | invalid | 0 | 0 |
| 45 | 30,10 | 37 | 30,13 | 0 | 0 | 0 | 0 | 0 | 0 | 0 | 0 | 0 | 0 | 0 |
| 41 | 30,13 | 41 | 30,14 | 1 | 0 | 0 | 0 | 0 | 0 | 0 | 0 | 0 | 0 | 0 |
| 37 | 30,54 | 45 | 31,19 | 0 | 0 | 0 | 0 | 0 | 0 | 0 | 0 | N/A | 0 | 0 |
| 43 | 31,05 | 48 | 31,19 | 0 | 0 | 0 | 0 | 0 | 0 | 0 | 0 | invalid | 0 | 0 |
| 46 | 31,54 | 46 | 31,34 | 1 | 0 | 0 | 0 | 0 | 0 | 0 | 0 | invalid | 0 | 0 |
| 48 | 32,06 | 43 | 31,61 | 0 | 0 | 0 | 0 | 0 | 0 | 0 | 0 | 0 | 0 | 0 |
| 47 | 35,19 | 47 | 34,55 | 0 | 0 | 0 | 0 | 0 | 0 | 0 | 0 | 0 | 0 | 0 |
| 49 | 35,22 | 50 | 35,83 | 0 | 0 | 0 | 0 | 0 | 0 | 0 | 0 | 1 | 0 | 0 |
| 50 | 36,36 | 49 | 36,04 | 0 | 0 | 0 | 0 | 0 | 0 | 0 | 0 | invalid | 0 | 0 |
