## Supplemental Table 2 for "Comparative sensitivity evaluation for 122 CE-marked SARS-CoV-2 antigen rapid tests"

Comparative evaluation results of SARS-CoV-2 antigen RDT missing the sensitivity criteria

(in alphabetical order of manufacturers)

| | | | | 1 | 2 | 3 | 4 | 5 | 6 | 7 | 8 | 9 | 10 | 11 | 12 | 13 | 14 | 15 | 16 | 17 |
|---|---|---|---|---|---|---|---|---|---|---|---|---|---|---|---|---|---|---|---|---|
| | | **Manufacturer** | | Acro Biotech Inc | Aikang Diagnostics Co., Ltd. | Beijing Savant Biotechnology Co., Ltd | Certest Biotec S. L. | Coris Bioconcept | Hangzhou AllTest Biotech Co. Ltd. | Hangzhou Biotest Biotech Co., Ltd. | Hangzhou Genesis Biocontrol Co., Ltd | Hangzhou Realy Tech Co., Ltd. | Inzek International Trading | Joinstar Biomedical Technology Co., Ltd | Joysbio (Tianjin) Biotechnology Co., Ltd. | Lionex GmbH | Medicon Co., Ltd. | Mexacare GmbH Heidelberg | nal von minden GmbH | Rapigen |
| | | **Test name** | | Acro COVID-19 Antigen Rapid Test | SARS-CoV-2 Antigen Test Kit (Immunochromatography) | New Coronavirus (SARS-CoV-2) N Protein Detection Kit (Fluorescence Immunchromatography) | CerTest SARS-CoV-2 | COVID-19 Ag Respi-Strip | COVID-19 AG AllTest | Lumiratek SARS-CoV-2 Antigen Rapid Test Cassette | KaiBiLi COVID-19 Antigen Rapid Test Device | Novel Coronavirus (SARS-Cov-2) Antigen Rapid Test Cassette (swab) | Biozek medical COVID-19 Antigen Rapid Test Cassette | COVID-19 Antigen Rapid Test (Latex) | Joysbio SARS-CoV-2 Antigen Rapid Test Kit (Colloidal Gold) | Lionex COVID-19 Ag Rapid Test | Trueline COVID-19 Ag Rapid Test | COVID-19 Antigen Schnelltest | dedicio Medical Test COVID-19 Ag plus Test | Biocredit COVID-19 Ag |
| **Panel 1 V1** | | **Panel 1 V2** | | Panel 1 V1 | Panel 1 V2 | Panel 1 V2 | Panel 1 V2 | Panel 1 V1 | Panel 1 V1 | Panel 1 V2 | Panel 1 V2 | Panel 1 V2 | Panel 1 V2 | Panel 1 V2 | Panel 1 V2 | Panel 1 V2 | Panel 1 V2 | Panel 1 V2 | Panel 1 V2 | Panel 1 V1 |
| **Pool Nr.** | **CT** | **Pool Nr.** | **CT** | | | | | | | | | | | | | | | | | |
| 1 | 17,55 | 1 | 17,31 | 1 | 1 | 0 | 1 | 1 | 1 | 1 | 1 | 1 | 1 | background | 1 | background | 1 | 1 | 1 | 1 |
| 6 | 20,20 | 2 | 19,08 | 0 | 1 | 0 | 1 | 1 | 0 | 1 | 1 | 1 | 1 | background | 1 | background | 1 | 1 | 1 | 0 |
| 5 | 20,28 | 3 | 19,62 | 0 | 0 | 0 | 0 | 1 | 0 | 1 | 1 | 1 | 1 | background | 1 | background | 1 | 1 | 1 | 1 |
| 3 | 20,38 | 5 | 20,60 | 0 | 0 | 0 | 1 | 1 | 1 | 1 | 1 | 1 | 1 | background | 1 | background | 1 | 1 | 1 | 1 |
| 2 | 20,54 | 4 | 20,61 | 0 | 0 | 0 | 1 | 1 | 1 | 1 | 1 | 1 | 1 | background | 1 | background | 1 | 1 | 1 | 0 |
| 4 | 20,98 | 6 | 21,21 | 0 | 0 | 0 | 1 | 1 | 0 | 0 | 1 | 1 | 1 | background | 1 | background | 1 | 1 | 0 | 0 |
| 7 | 21,71 | 12 | 22,12 | 0 | 0 | 0 | 0 | 0 | 0 | 0 | 1 | 1 | 0 | background | 1 | background | 0 | 0 | 0 | 0 |
| 12 | 21,82 | 7 | 22,15 | 0 | 0 | 0 | 0 | 0 | 0 | 0 | 0 | 1 | 1 | background | 0 | background | 1 | 1 | 0 | 0 |
| 8 | 21,95 | 8 | 22,32 | 0 | 0 | 0 | 0 | 0 | 0 | 0 | 1 | 0 | 1 | background | 1 | background | 0 | 0 | 0 | 0 |
| 9 | 22,14 | 16 | 22,88 | 1 | 0 | 0 | 0 | 0 | 0 | 0 | 1 | 0 | 0 | background | 0 | background | 1 | 0 | 1 | 0 |
| 11 | 22,34 | 9 | 23,13 | 0 | 0 | 0 | 0 | 0 | 0 | 0 | 0 | 1 | 1 | background | 0 | background | 1 | 1 | 0 | 0 |
| 16 | 22,55 | 11 | 23,13 | 0 | 0 | 0 | 0 | 0 | 0 | 0 | 0 | 0 | 0 | background | 0 | background | 1 | 0 | 0 | 0 |
| 10 | 22,88 | 10 | 23,21 | 1 | 0 | 0 | 0 | 0 | 0 | 0 | 0 | 1 | 0 | background | 0 | background | 0 | 0 | 0 | 0 |
| 13 | 23,32 | 15 | 24,38 | 0 | 0 | 0 | 0 | 0 | 0 | 0 | 0 | 0 | 0 | background | 0 | background | 0 | 0 | 0 | 0 |
| 17 | 24,00 | 23 | 24,45 | 0 | 0 | 0 | 0 | 0 | 0 | 0 | 0 | 0 | 0 | background | 0 | background | 0 | 0 | 0 | 0 |
| 23 | 24,04 | 17 | 24,81 | 0 | 0 | 0 | 0 | 0 | 0 | 0 | 0 | 0 | 0 | background | 0 | background | 0 | 0 | 0 | 0 |
| 15 | 24,14 | 14 | 24,97 | 0 | 0 | 0 | 0 | 0 | 0 | 0 | 0 | 0 | 0 | background | 0 | background | 0 | 1 | 0 | 0 |
| 14 | 24,28 | 25 | 25,07 | 0 | 0 | 0 | 0 | 0 | 0 | 0 | 0 | 0 | 0 | background | 0 | background | 1 | 0 | 0 | 0 |
| 27 | 25,14 | 24 | 25,20 | 0 | 0 | 0 | 0 | 0 | 0 | 0 | 0 | 0 | 0 | background | 0 | background | 0 | 0 | 0 | 0 |
| 24 | 25,24 | 13 | 25,29 | 0 | 0 | 0 | 0 | 0 | 0 | 0 | 0 | 0 | 0 | background | 0 | background | 0 | 0 | 0 | 0 |
| 31 | 25,27 | 19 | 25,45 | 0 | 0 | 0 | 0 | 0 | 0 | 0 | 0 | 0 | 0 | background | 0 | background | 0 | 0 | 0 | 0 |
| 18 | 25,30 | 21 | 25,95 | 0 | 0 | 0 | 0 | 0 | 0 | 0 | 0 | 0 | 0 | background | 0 | background | 0 | 0 | 0 | 0 |
| 26 | 25,47 | 27 | 26,12 | 0 | 0 | 0 | 0 | 0 | 0 | 0 | 0 | 0 | 0 | background | 0 | background | 0 | 0 | 0 | 0 |
| 19 | 25,50 | 31 | 26,24 | 0 | 0 | 0 | 0 | 0 | 0 | 0 | 0 | 0 | 0 | background | 1 | background | 0 | 0 | 0 | 0 |
| 21 | 25,54 | 26 | 26,32 | 0 | 0 | 0 | 0 | 0 | 0 | 0 | 0 | 0 | 0 | background | 0 | background | 0 | 0 | 0 | 0 |
| 22 | 25,87 | 32 | 26,64 | 0 | 0 | 0 | 0 | 0 | 0 | 0 | 0 | 0 | 0 | background | 0 | background | 0 | 0 | 0 | 0 |
| 20 | 26,27 | 35 | 26,66 | 0 | 0 | 0 | 0 | 0 | 0 | 0 | 0 | 0 | 0 | background | 0 | background | 0 | 0 | 0 | 0 |
| 32 | 26,44 | 36 | 27,05 | 0 | 0 | 0 | 0 | 0 | 0 | 0 | 0 | 0 | 0 | background | 0 | background | 0 | 0 | 0 | 0 |
| 35 | 27,04 | 30 | 27,24 | 0 | 0 | 0 | 0 | 0 | 0 | 0 | 0 | 0 | 0 | background | 0 | background | 0 | 0 | 0 | 0 |
| 28 | 27,14 | 29 | 27,34 | 0 | 0 | 0 | 0 | 0 | 0 | 0 | 0 | 0 | 0 | background | 0 | background | 0 | 0 | 0 | 0 |
| 29 | 27,15 | 28 | 27,41 | 0 | 0 | 0 | 0 | 0 | 0 | 0 | 0 | 0 | 0 | background | 0 | background | 0 | 0 | 0 | 0 |
| 40 | 27,65 | 22 | 27,42 | 0 | 0 | 0 | 0 | 0 | 0 | 0 | 0 | 0 | 0 | background | 0 | background | 0 | 0 | 0 | 0 |
| 34 | 27,89 | 34 | 27,82 | 0 | 0 | 0 | 0 | 0 | 0 | 0 | 0 | 0 | 0 | background | 0 | background | 0 | 0 | 0 | 0 |
| 36 | 28,13 | 40 | 28,19 | 0 | 0 | 0 | 0 | 0 | 0 | 0 | 0 | 0 | 0 | background | 0 | background | 0 | 0 | 0 | 0 |
| 38 | 28,14 | 18 | 28,33 | 0 | 0 | 0 | 0 | 0 | 0 | 0 | 0 | 0 | 0 | background | 0 | background | 0 | 0 | 0 | 0 |
| 42 | 28,43 | 33 | 28,92 | 0 | 0 | 0 | 0 | 0 | 0 | 0 | 0 | 0 | 0 | background | 0 | background | 0 | 0 | 0 | 0 |
| 30 | 28,86 | 38 | 29,36 | 0 | 0 | 0 | 0 | 0 | 0 | 0 | 0 | 0 | 0 | background | 0 | background | 0 | 0 | 0 | 0 |
| 33 | 28,96 | 20 | 29,46 | 0 | 0 | 0 | 0 | 0 | 0 | 0 | 0 | 0 | 0 | background | 0 | background | 0 | 0 | 0 | 0 |
| 44 | 29,24 | 42 | 29,48 | 0 | 0 | 0 | 0 | 0 | 0 | 0 | 0 | 0 | 0 | background | 0 | background | 0 | 0 | 0 | 0 |
| 25 | 29,70 | 44 | 29,51 | 0 | 0 | 0 | 0 | 0 | 0 | 0 | 0 | 0 | 0 | background | 0 | background | 0 | 1 | 0 | 0 |
| 39 | 29,76 | 39 | 30,12 | 0 | 0 | 0 | 0 | 0 | 0 | 0 | 0 | 0 | 0 | background | 0 | background | 0 | 0 | 0 | 0 |
| 45 | 30,10 | 37 | 30,13 | 0 | 0 | 0 | 0 | 0 | 0 | 0 | 0 | 0 | 0 | background | 0 | background | 0 | 0 | 0 | 0 |
| 41 | 30,13 | 41 | 30,14 | 0 | 0 | 0 | 0 | 0 | 0 | 0 | 0 | 0 | 0 | background | 0 | background | 0 | 0 | 0 | 0 |
| 37 | 30,54 | 45 | 31,19 | 0 | 0 | 0 | 0 | 0 | 0 | 0 | 0 | 0 | 0 | background | 0 | background | 0 | 0 | 0 | 0 |
| 43 | 31,05 | 48 | 31,19 | 0 | 0 | 0 | 0 | 0 | 0 | 0 | 0 | 0 | 0 | background | 0 | background | 0 | 0 | 0 | 0 |
| 46 | 31,54 | 46 | 31,34 | 0 | 0 | 0 | 0 | 0 | 0 | 0 | 0 | 0 | 0 | background | 0 | background | 0 | 0 | 0 | 0 |
| 48 | 32,06 | 43 | 31,61 | 0 | 0 | 0 | 0 | 0 | 0 | 0 | 0 | 0 | 0 | background | 0 | background | 0 | 0 | 0 | 0 |
| 47 | 35,19 | 47 | 34,55 | 0 | 0 | 0 | 0 | 0 | 0 | 0 | 0 | 0 | 0 | background | 0 | background | 0 | 0 | 0 | 0 |
| 49 | 35,22 | 50 | 35,83 | 0 | 0 | 0 | 0 | 0 | 0 | 0 | 0 | 0 | 0 | background | 0 | background | 0 | 0 | 0 | 0 |
| 50 | 36,36 | 49 | 36,04 | 0 | 0 | 0 | 0 | 0 | 0 | 0 | 0 | 0 | 0 | background | 0 | background | 0 | 0 | 0 | 0 |

Supplemental Table 2

Comparative evaluation results of SARS-CoV-2 antigen RDT missing the sensitivity criteria

(in alphabetical order of manufacturers)

| | | | | 18 | 19 | 20 | 21 | 22 | 23 | 24 | 25 | 26 |
|---|---|---|---|---|---|---|---|---|---|---|---|---|
| | | **Manufacturer** | | Servoprax | Spring Healthcare Services SP zoo | SureScreen Diagnostics Ltd | TaiDoc Technology Corp. | Unioninvest | VivaChek Biotech (Hangzhou) Co,Ltd. | VivaChek Biotech (Hangzhou) Co,Ltd. | W.H.P.M, Inc | Xiamen Zhongsheng Langjie Biotechnology Co., Ltd |
| | | **Test name** | | Cleartest Coronaantigen | SARS-Cov-2 Antigen Rapid Test Cassette (swab) | COVID-19 Antigen Rapid Test Cassette | FORA COVID-19 ANTIGEN RAPID TEST | Unibioscience COVID-19 Rapid Antigen Test | VivaDiag SARS-CoV-2 Ag Rapid Test | VivaDiag Pro SARS-CoV-2 Ag Rapid Test | First SIGN SARS-CoV-2 Antigen Test | Covid-19 Antigen Test Cassette |
| **Panel 1 V1** | | **Panel 1 V2** | | Panel 1 V1 | Panel 1 V2 | Panel 1 V2 | Panel 1 V1 | Panel 1 V1 | Panel 1 V1 | Panel 1 V2 | Panel 1 V2 | Panel 1 V2 |
| **Pool Nr.** | **CT** | **Pool Nr.** | **CT** | | | | | | | | | |
| 1 | 17,55 | 1 | 17,31 | 1 | 1 | 1 | 1 | 0 | 1 | 1 | 1 | 1 |
| 6 | 20,20 | 2 | 19,08 | 1 | 1 | 1 | 1 | 0 | 1 | 1 | 1 | 1 |
| 5 | 20,28 | 3 | 19,62 | 1 | 1 | 1 | 1 | 0 | 1 | 1 | 1 | 0 |
| 3 | 20,38 | 5 | 20,60 | 1 | 1 | 1 | 1 | 0 | 1 | 1 | 1 | 0 |
| 2 | 20,54 | 4 | 20,61 | 1 | 1 | 1 | 1 | 0 | 1 | 1 | 1 | 0 |
| 4 | 20,98 | 6 | 21,21 | 1 | 0 | 1 | 0 | 0 | 1 | 1 | 1 | 0 |
| 7 | 21,71 | 12 | 22,12 | 1 | 0 | 1 | 0 | 0 | 1 | 1 | 0 | 0 |
| 12 | 21,82 | 7 | 22,15 | 1 | 0 | 1 | 0 | 0 | 1 | 1 | 0 | 0 |
| 8 | 21,95 | 8 | 22,32 | 1 | 0 | 1 | 0 | 0 | 0 | 1 | 1 | 0 |
| 9 | 22,14 | 16 | 22,88 | 1 | 0 | 0 | 0 | 0 | 1 | 0 | 0 | 0 |
| 11 | 22,34 | 9 | 23,13 | 0 | 0 | 0 | 0 | 0 | 0 | 1 | 1 | 0 |
| 16 | 22,55 | 11 | 23,13 | 0 | 0 | 0 | 0 | 0 | 0 | 0 | 0 | 0 |
| 10 | 22,88 | 10 | 23,21 | 1 | 0 | 0 | 0 | 0 | 0 | 1 | 0 | 0 |
| 13 | 23,32 | 15 | 24,38 | 1 | 0 | 0 | 0 | 0 | 0 | 0 | 0 | 0 |
| 17 | 24,00 | 23 | 24,45 | 0 | 0 | 0 | 0 | 0 | 0 | 0 | 0 | 0 |
| 23 | 24,04 | 17 | 24,81 | 0 | 0 | 0 | 0 | 0 | 0 | 0 | 0 | 0 |
| 15 | 24,14 | 14 | 24,97 | 0 | 0 | 0 | 0 | 0 | 0 | 0 | 0 | 0 |
| 14 | 24,28 | 25 | 25,07 | 0 | 0 | 0 | 0 | 0 | 0 | 0 | 0 | 0 |
| 27 | 25,14 | 24 | 25,20 | 0 | 0 | 0 | 0 | 0 | 0 | 0 | 0 | 0 |
| 24 | 25,24 | 13 | 25,29 | 0 | 0 | 0 | 0 | 0 | 0 | 0 | 0 | 0 |
| 31 | 25,27 | 19 | 25,45 | 0 | 0 | 0 | 0 | 0 | 0 | 0 | 0 | 0 |
| 18 | 25,30 | 21 | 25,95 | 0 | 0 | 0 | 0 | 0 | 0 | 0 | 0 | 0 |
| 26 | 25,47 | 27 | 26,12 | 0 | 0 | 0 | 0 | 0 | 0 | 0 | 0 | 0 |
| 19 | 25,50 | 31 | 26,24 | 0 | 0 | 0 | 0 | 0 | 0 | 0 | 0 | 0 |
| 21 | 25,54 | 26 | 26,32 | 0 | 0 | 0 | 0 | 0 | 0 | 0 | 0 | 0 |
| 22 | 25,87 | 32 | 26,64 | 0 | 0 | 0 | 0 | 0 | 0 | 0 | 0 | 0 |
| 20 | 26,27 | 35 | 26,66 | 0 | 0 | 0 | 0 | 0 | 0 | 0 | 0 | 0 |
| 32 | 26,44 | 36 | 27,05 | 0 | 0 | 0 | 0 | 0 | 0 | 0 | 0 | 0 |
| 35 | 27,04 | 30 | 27,24 | 0 | 0 | 0 | 0 | 0 | 0 | 0 | 0 | 0 |
| 28 | 27,14 | 29 | 27,34 | 0 | 0 | 0 | 0 | 0 | 0 | 0 | 0 | 0 |
| 29 | 27,15 | 28 | 27,41 | 0 | 0 | 0 | 0 | 0 | 0 | 0 | 0 | 0 |
| 40 | 27,65 | 22 | 27,42 | 0 | 0 | 0 | 0 | 0 | 0 | 0 | 0 | 0 |
| 34 | 27,89 | 34 | 27,82 | 0 | 0 | 0 | 0 | 0 | 0 | 0 | 0 | 0 |
| 36 | 28,13 | 40 | 28,19 | 0 | 0 | 0 | 0 | 0 | 0 | 0 | 0 | 0 |
| 38 | 28,14 | 18 | 28,33 | 0 | 0 | 0 | 0 | 0 | 0 | 0 | 0 | 0 |
| 42 | 28,43 | 33 | 28,92 | 0 | 0 | 0 | 0 | 0 | 0 | 0 | 0 | 0 |
| 30 | 28,86 | 38 | 29,36 | 0 | 0 | 0 | 0 | 0 | 0 | 0 | 0 | 0 |
| 33 | 28,96 | 20 | 29,46 | 0 | 0 | 0 | 0 | 0 | 0 | 0 | 0 | 0 |
| 44 | 29,24 | 42 | 29,48 | 0 | 0 | 0 | 0 | 0 | 0 | 0 | 0 | 0 |
| 25 | 29,70 | 44 | 29,51 | 0 | 0 | 0 | 0 | 0 | 0 | 0 | 0 | 0 |
| 39 | 29,76 | 39 | 30,12 | 0 | 0 | 0 | 0 | 0 | 0 | 0 | 0 | 0 |
| 45 | 30,10 | 37 | 30,13 | 0 | 0 | 0 | 0 | 0 | 0 | 0 | 0 | 0 |
| 41 | 30,13 | 41 | 30,14 | 0 | 0 | 0 | 0 | 0 | 0 | 0 | 0 | 0 |
| 37 | 30,54 | 45 | 31,19 | 0 | 0 | 0 | 0 | 0 | 0 | 0 | 0 | 0 |
| 43 | 31,05 | 48 | 31,19 | 0 | 0 | 0 | 0 | 0 | 0 | 0 | 0 | 0 |
| 46 | 31,54 | 46 | 31,34 | 0 | 0 | 0 | 0 | 0 | 0 | 0 | 0 | 0 |
| 48 | 32,06 | 43 | 31,61 | 0 | 0 | 0 | 0 | 0 | 0 | 0 | 0 | 0 |
| 47 | 35,19 | 47 | 34,55 | 0 | 0 | 0 | 0 | 0 | 0 | 0 | 0 | 0 |
| 49 | 35,22 | 50 | 35,83 | 0 | 0 | 0 | 0 | 0 | 0 | 0 | 0 | 0 |
| 50 | 36,36 | 49 | 36,04 | 0 | 0 | 0 | 0 | 0 | 0 | 0 | 0 | 0 |
